# Segmental analysis in cervical spinal cord injury reveals the recovery potential of hand muscles with preserved corticospinal tract: Insights beyond impairment scales

**DOI:** 10.1101/2021.11.30.21265051

**Authors:** Gustavo Balbinot, Guijin Li, Sukhvinder Kalsi-Ryan, Rainer Abel, Doris Maier, Yorck-Bernhard Kalke, Norbert Weidner, Rüdiger Rupp, Martin Schubert, Armin Curt, Jose Zariffa

## Abstract

Cervical spinal cord injury (SCI) severely impacts widespread bodily functions with extensive impairments for individuals, who prioritize regaining hand function. Although prior work has focused on the recovery at the person-level, the factors determining the recovery potential of individual muscles are poorly understood. There is a need for changing this paradigm in the field by moving beyond person-level classification of residual strength and sacral sparing to a muscle-specific analysis with a focus on the role of corticospinal tract (CST) sparing. The most striking part of human evolution involved the development of dextrous hand use with a respective expansion of the sensorimotor cortex controlling hand movements, which, because of the extensive CST projections, may constitute a drawback after SCI. Here, we investigated the muscle-specific natural recovery after cervical SCI in 748 patients from the European Multicenter Study about SCI (EMSCI), one of the largest datasets analysed to date. All participants were assessed within the first 4 weeks after SCI and re-assessed at 12, 24, and 48 weeks. Subsets of individuals underwent electrophysiological multimodal evaluations to discern CST and lower motor neuron (LMN) integrity [motor evoked potentials (MEP): N = 203; somatosensory evoked potentials (SSEP): N = 313; nerve conduction studies (NCS): N = 280]. We show the first evidence of the importance of CST sparing for proportional recovery in SCI, which is known in stroke survivors to represent the biological limits of structural and functional plasticity. In AIS D, baseline strength is a good predictor of segmental muscle strength recovery, while the proportionality in relation to baseline strength is lower for AIS B/C and breaks for AIS A. More severely impaired individuals showed non-linear and more variable recovery profiles, especially for hand muscles, while measures of CST sparing (by means of MEP) improved the prediction of hand muscle strength recovery. Therefore, assessment strategies for muscle-specific motor recovery in acute SCI improve by accounting for CST sparing and complement gross person-level predictions. The latter is of paramount importance for clinical trial outcomes and to target neurorehabilitation of upper limb function, where any single muscle function impacts the outcome of independence in cervical SCI.

**Graphical abstract:** 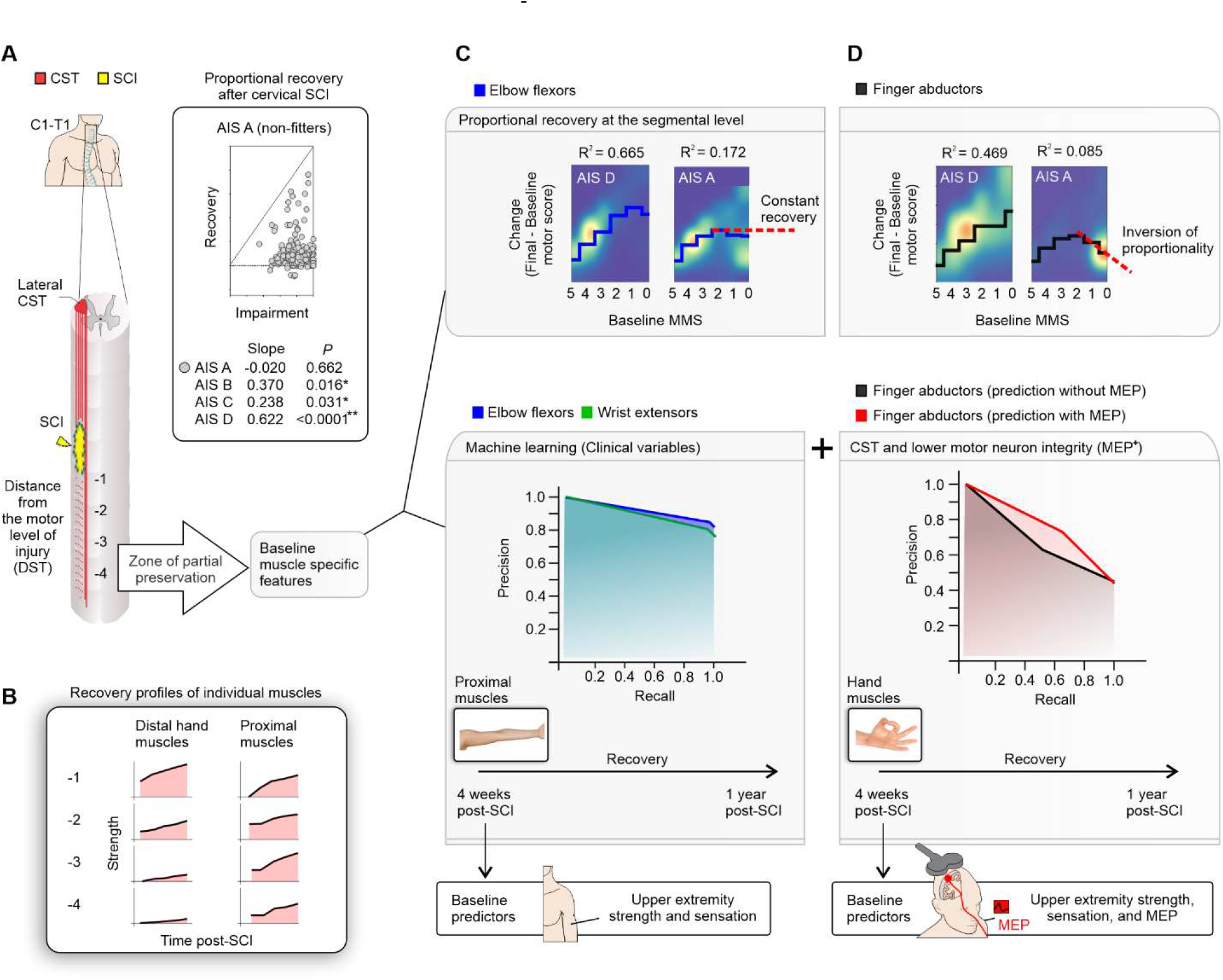

Segmental analysis in cervical spinal cord injury reveals the recovery potential of hand muscles with preserved corticospinal tract: Insights beyond impairment scales. (A, upper panels) Cervical SCI (yellow) may cause impairment of motor function below the level of lesion depending on the completeness of the injury. Individuals with a sensorimotor complete lesion (AIS A), as defined by the absence of sacral sparing, show a non-proportional strength recovery as related to the baseline strength, in contrast to less severely affected patients (AIS B-D) - reflecting the limitations on structural and functional plasticity in this group. (A, lower panel) The area of spinal cord damage typically extents across several segments below the level of lesion with variable preservation of muscle innervation and is described as zone of partial preservation (ZPP). (B) The recovery of hand muscle strength is more challenged compared to more proximal muscles when accounting for the distance from the level of lesion. (C, upper panel) The strength recovery of proximal muscles is proportional to the baseline strength in AIS D (great R^2^ values) but limited in AIS A, likely indicating the limits of recovery in severe SCI. (C, lower panel) Also, additional clinical baseline variables [e.g., distance from the motor level of injury (DST); pin prick (PP) and light touch (LT) sensation] primarily increased the prediction of strength recovery for proximal muscles, becoming less effective in more distal muscles, such as the intrinsic hand muscles. (D) Overall, the proportional prediction of strength recovery in distal hand muscles is less strong while failing in AIS A (inversion of proportionality). (D, lower panel) The addition of neurophysiological baseline measures related to CST integrity (by means of MEP) increased the prediction of strength recovery of hand muscles, indicating the importance of residual CST projections to spinal motoneurons for hand strength recovery. Clinical studies aiming at restitution of hand function after SCI may benefit from the addition of MEP assessments early after the SCI, to unveil hand muscles with a potential for recovery.

## Introduction

Spinal cord injury (SCI) is defined as damage to the spinal cord resulting in temporary or permanent changes in its function.^1^ SCI poses important physical and social consequences for the affected individuals, and the care of individuals with SCI requires substantial efforts. The development of effective treatments becomes crucially important to enhance spinal cord function, and subsequently, improve sensorimotor function and minimize secondary complications. Understanding sensorimotor recovery under the current standard of care enables the identification and prediction of persistent functional impairments that may be improved by treatment, and is essential to accurately assess the impact of interventions. In tetraplegia, the improvement in upper limb motor function is important and regaining hand function is considered a high priority.^2^ Although extensive effort has been devoted to understanding recovery of upper extremity function and strength^3–12^, little is known about how the segmental innervation of upper limb muscles recovers after SCI. ^13^ It is known that the impairment of upper limb muscles is related to task performance^8, 14^ and assessing strength of upper extremity muscles enables to predict upper limb function. ^15^ Nonetheless, the specific recovery profile of single upper limb muscles is still poorly understood, especially that of the hand muscles.^13^

Several factors may contribute to variations in recovery profiles across upper limb muscles. Upper limb muscles are controlled by integrated and relatively overlapped representations in the motor cortex^16^, but the cortical representation of hand muscles is larger, with extensive corticospinal tract (CST) connections to cervical spinal motoneurons.^17–19^ It is thought that spinal motoneurons of the distal compared to proximal upper limb muscles receive greater input from the primary motor cortex through the CST to execute more refined, versatile fine movements.^20–22^ When the spinal cord is injured, motor tracts may be damaged affecting the integrity of the CST.^23–27^ The role of CST integrity on upper limb motor recovery is a topic extensively studied in stroke. The relationship between motor recovery and the initial impairment reflects the biological limits of structural and functional plasticity. Individuals with a stroke severely affecting CST integrity display great impairment and limited recovery of upper limb function and do not fit the proportional relationship that has been observed between the amount of recovery and the initial impairment in individuals with less CST damage.^28, 29^ It is known that ‘non-fitters’ have limited performance in tasks related to wrist/hand dexterity, which is also indicative of a more pronounced CST disruption.^30^ In SCI, the lesion will often affect the projections from the CST and other descending tracts to spinal motoneurons (i.e., axonal lesions of upper motor neurons; UMN) and/or directly damage α-motoneurons [i.e., lower motor neuron (LMN) lesion], depending on the extent, location, and severity of the lesion. UMN versus LMN damage is not distinguished by clinical exam (e.g., International Standards for Neurological Classification of Spinal Cord Injury; ISNCSCI).^31^ Thereby, the effect of UMN and LMN lesions may vary across upper limb muscles; for anatomical reasons CST damage may have more pronounced effects on distal movements, while LMN damage is likely to be more pronounced at or immediately below the lesion. In addition, the upper limb is comprised of muscles specialized for both gross and fine motor function, leading to variations in the number and size of motor units and muscle fiber types across muscles.^32, 33^ The impact of such variations on muscle functional recovery in SCI is poorly understood.

Established recovery profiles after SCI have not distinguished the development of spastic or flaccid muscles weakness, and summed motor scores (UEMS) do not discern the recovery of distal or proximal upper limb muscles. Although it is known that the residual muscle strength early after SCI is indicative of preserved CST connections and a good predictor of summed upper limb strength recovery^6, 7^, the prediction of individual myotomes is lacking. Neurophysiological assessments such as motor evoked potentials (MEPs), somatosensory evoked potentials (SSEPs) and compound muscle action potential (CMAPs) have been applied to assess CST and/or α- motoneuron integrity and the natural extent of spinal neural recovery contributing to the prediction of gross functions like walking and independence^4^, and may be beneficial as well for predicting myotome recovery. Here, the muscle-specific approach supports an emerging scenario in the field aimed at better understanding motor discomplete lesions^34–37^ and the importance of lateral tract sparing for recovery prognostics.^38, 39^

The goal of the present study is to explore if segmental innervation as assessed in single upper limb muscles exhibits different strength recovery profiles after cervical SCI. We aim to identify factors predictive of segmental strength recovery in upper extremity muscles and hypothesize that neuroanatomical factors as related to segmental muscles (i.e., extent of corticospinal connections, and distance to the motor level of lesion) may affect the potential for recovery.

## Materials and methods

### Study design

The study is based on the European Multicenter Study about SCI (EMSCI; ClinicalTrials.gov Identifier: NCT01571531) investigating the natural recovery after SCI. The inclusion criteria of EMSCI are: (1) single event traumatic or ischemic para- or tetraplegia, (2) first assessment possible within the first 4 weeks after incidence, (3) patient capable and willing of giving informed consent. Seven hundred and ninety-nine participants with cervical SCI were enrolled in dedicated SCI centres: the Hohe Warte Bayreuth (Bayreuth, Germany), BG-Trauma Center (Murnau, Germany), RKU Universitäts- und Rehabilitationskliniken Ulm (Ulm, Germany), Spinal Cord Injury Center of Heidelberg University Hospital (Heidelberg, Germany), and Spinal Cord Injury Center - Balgrist University Hospital (Zurich, Switzerland). The research followed the Declaration of Helsinki and was approved by the Institutional Review Board of the abovementioned institutions: Bayrische Landesärztekammer, Ethik-Kommission (REB #188/2003; Bayreuth, Germany), Ethik-Kommission der Bayerischen Landesärztekammer (REB approval was waived because the project was treated as a data registry, but informed consent was obtained from all participants; Murnau, Germany), Universität Ulm Ethikkommission (REB #71/2005; Ulm, Germany), Universität Heidelberg Ethikkommission der Med. Fakultät (REB #S-188/2003; Heidelberg, Germany), Kanton Zürich Kantonale Ethikkommission (REB #EK-03/2004/PB_2016- 00293; Zurich, Switzerland). Fifty-one individuals were excluded because of incomplete ISNCSCI assessments at baseline, as such, data from 748 individuals were analysed. All participants were assessed within the first 4 weeks (Mean = 31 days; SD = 6.8 days) after SCI and re-assessed at 12 (Mean = 84.6 days; SD = 8.5 days), 24 (Mean = 168.5 days; SD = 11.1 days), and 48 weeks (Mean = 356.7 days; SD = 54 days). A subset of participants (N = 440) was additionally assessed in the very acute phase of SCI (Mean = 8.7 days; SD = 4.6 days), which was specifically used to describe motor recovery. Subsets of participants also underwent electrophysiological multimodal assessments of motor evoked potentials (MEPs; N = 203), somatosensory evoked potentials (SSEPs; N = 313), and nerve conduction studies (NCS; N = 280) of the nerves of the upper limb innervating hand muscles. The research followed the Declaration of Helsinki and was approved by the Institutional Review Board of the above-mentioned institutions. Supplementary Table 1 shows demographic and clinical characteristics of the research participants.

Muscle strength was measured according to ISNCSCI in ten key muscles for each side of the body for each participant: five upper limb muscles [elbow flexors (C5), wrist extensors (C6), elbow extensors (C7), finger flexors (C7), and finger abductors (T1)] and five lower limb muscles [hip flexors (L2), knee extensors (L3), ankle dorsiflexors (L4), long toe extensors (L5) and ankle plantar flexors (S1)]. Each muscle was scored from 0-5 (Muscle Motor Score; MMS) following the recommendations of the ISNCSCI.^31, 40^ Thus, the total motor score has a maximum of 50 per side, and 100 per person. The upper extremity motor score (UEMS) consists of a maximum of 25 per side and 50 per person.

In a first step, we described the segmental strength recovery profiles after cervical SCI. In a second step, we investigated the ability of several baseline anatomical and injury characteristics (summarized in Figure 1) to predict segmental recovery.

**Figure 1.**
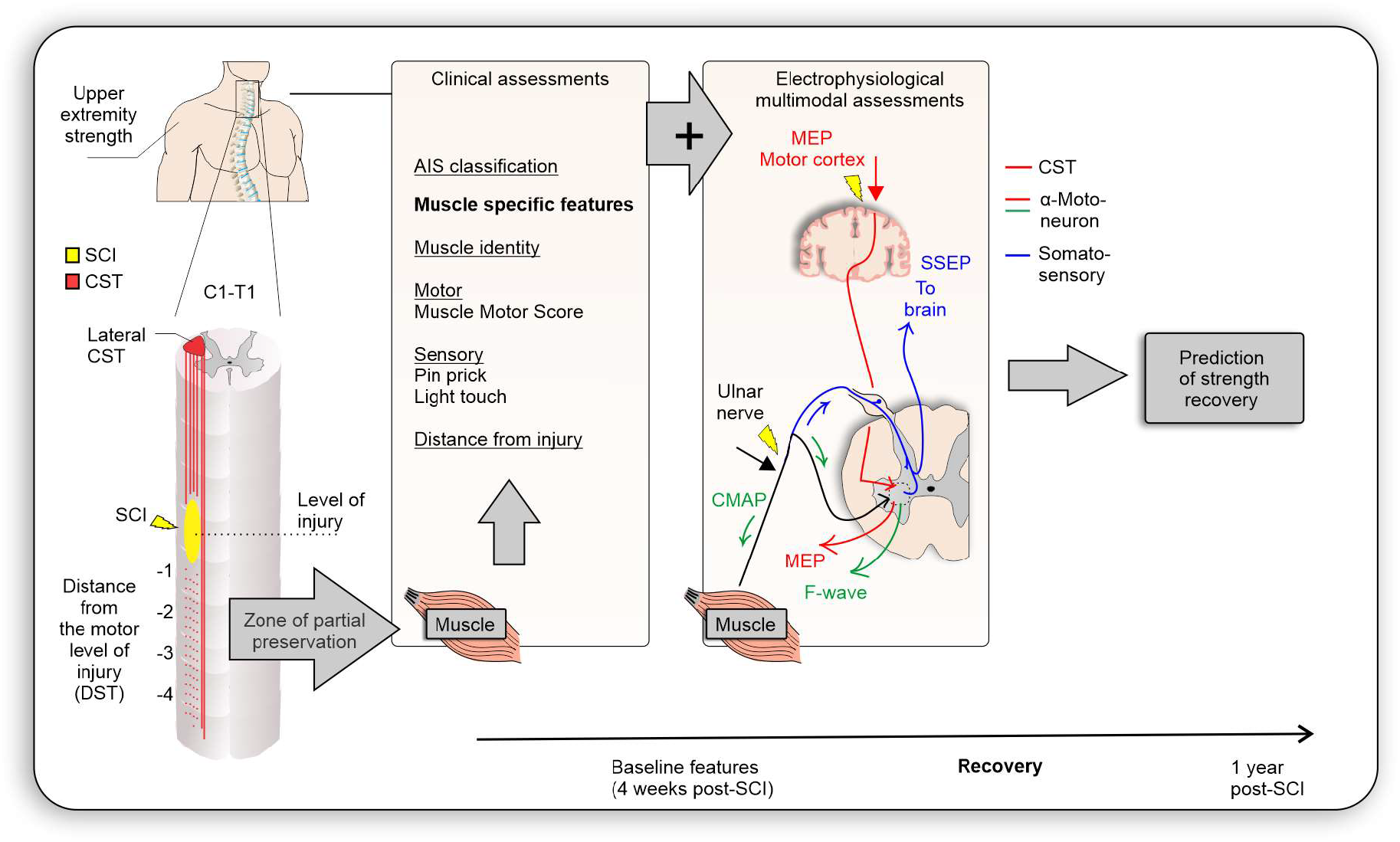
Anatomical and injury characteristics hypothesized to be determinants of segmental muscle recovery. Muscle identity may play a role due to variations in cortical representation and CST projections. Remaining spinal innervation after injury may be reflected in the muscle motor score, sensory scores in corresponding dermatomes, and electrophysiological assessments (MEP, SSEP). NCS may improve predictions by providing information about α- motoneuron damage. AIS grade and distance from injury further determine the capacity for neurorecovery.

### Segmental strength recovery of upper limb muscles after SCI

Non-parametric statistics and non-linear regression using random forest regressors were used to explore segmental strength recovery after SCI. The distance (DST) in myotomes between the ISNCSCI motor level and the muscle myotome on each side of the body was used to split the dataset, in order to control for the distance from the motor level when comparing muscles (negative DST values denote myotomes caudal to the motor level). It is known that the recovery of motor function in spinal segments below the ISNCSCI motor level will typically occur approximately one to three levels caudal to it (DSTs -1 to -3).^3^ Based on this information, our analysis encompassed muscles 1 to 4 levels below the motor level (DSTs -1 to -4). Muscles from the left and right sides of the same participant were considered as independent samples after correcting for DST. For the random forest non-linear regression and classification models, muscles with a baseline strength of 5 were excluded to control for ceiling effects.

### Prediction of segmental strength recovery

#### Prediction of strength recovery after cervical SCI: the role of baseline MMS

The residual strength after SCI is indicative of preserved supraspinal connections to the muscles. To test if strength recovery is related to the amount of residual strength at baseline, we assessed the recovery using the proportional recovery framework previously employed to describe stroke recovery.^28–30, 41–48^ The change of MMS between baseline (4 weeks) and endpoint (48 weeks) were regressed against baseline MMS, the initially preserved motor function, to predict motor recovery in relation to the initial impairment.

#### Prediction of strength recovery after cervical SCI: the role of additional segment-specific variables

In addition to the baseline MMS, we explored the inclusion of variables extracted from the sensory components of the ISNCSCI as additional features in the machine learning models.^31^ The light touch (LT) and pin prick (PP) sensation scores of the dermatome corresponding to each myotome of the upper extremity were analyzed. In addition to LT and PP scores, the DST was also considered as a feature in the machine learning models. General models included all AIS grades and muscles (features used in model 1: AIS, MMS, DST, LT, and PP). Muscle identity (i.e., key muscles in the ISNCSCI: C5/elbow flexors, C6/wrist extensors, C7/elbow extensors, C8/finger flexors, or T1/finger abductors) was taken into account in a subsequent step (features used in model 1 + Muscle identity: AIS, Muscle, MMS, DST, LT, and PP). We also created muscle-specific models by using data from each individual key muscle (models 2-6) rather than pooled data from all muscles. These models were important to understand how the strength recovery prediction differed between muscles. Random forest classifiers were used to predict segmental strength recovery after SCI: model 2 (elbow flexors), model 3 (wrist extensors), model 4 (elbow extensors), model 5 (finger flexors), and model 6 (finger abductors).

#### Prediction of strength recovery after cervical SCI: the role of electrophysiological multimodal assessments

MMS alone may not reflect CST and LMN sparing^49^ and, therefore, we explored the predictive value of electrophysiological multimodal assessments in a subsample of the participants. This subsample consisted of individuals classified as AIS A/B/C that had undergone electrophysiological multimodal assessments at baseline [within the first 4 weeks (Mean = 31 days; SD = 6.8 days)]. The assessments conducted were motor evoked potentials (MEPs) on the abductor digiti minimi, somatosensory evoked potentials (SSEPs) from the ulnar nerve, and nerve conduction studies (NCS) of the ulnar nerve. A detailed description of the materials and methods used in the electrophysiological multimodal assessments of the hand muscles is presented in the Supplementary Material.^4, 23, 27^

##### Electrophysiological multimodal assessments scoring system

Transformation to a scoring system was guided by clinical normative values.^4^ All neurophysiological examinations were rated as normal (2 points), impaired (1 point), or abolished (0 points) as shown in Supplementary Table 2. The scoring system resulted in an ordinal value with a maximum of 3 points for MEP (MEP score), 3 points for SSEP (SSEP score), and 3 points for NCS (NCS score).

### Time-course of motor evoked and compound muscle action potentials recovery

For some participants, follow-up assessments of MEP were performed. The resulting time-course of MEP amplitude recovery provides an electrophysiological perspective on motor recovery. Muscles at and up to eight segments caudal to the motor level of SCI were considered for the analysis of the time-course of MEP recovery. Muscles with a baseline MMS of 5 were excluded to reduce ceiling effects. The follow-up assessment was conducted at different time points. A total of 259 abductor digiti minimi muscles MEPs were available at baseline, but in 46 only the baseline MEP assessment was performed and those were not included in the analysis. A total of 213 MEPs were included in this analysis, of which 133 had the endpoint MEP assessment conducted at 48 weeks post-SCI, 16 at 24 weeks, and 64 at 12 weeks. A total of 332 abductor digiti minimi muscles CMAPs were available at baseline, but in 27 muscles only the baseline CMAP assessment was performed and those were not included in the analysis. A total of 305 CMAPs were included in this analysis, of which 196 had the endpoint CMAP assessment conducted at 48 weeks post-SCI, 19 at 24 weeks, and 90 at 12 weeks.

### Statistical analysis

Statistical significance was set at α = 0.05. The analysis was conducted using Python scikit-learn (machine learning analysis and data visualization), SPSS^®^ Statistics (descriptive analysis, median comparisons), Excel (data sorting), LabVIEW^®^ (data visualization and sorting), and GraphPad Prism^®^ (data visualization and descriptive analysis).

### Descriptive and median comparisons

Data normality was assessed using the Shapiro-Wilk test. The unit of measure was each muscle and data were expressed using median or median, interquartile intervals, and 5-95 percentiles, unless otherwise noted. The McNemar’s test was used to compare muscle strength recovery (%muscles with MMS ≥ 3) over time. The Kruskal-Wallis H test was used as a non-parametric alternative to the one-way ANOVA to determine if there were statistically significant differences between the strength of distinct ISNCSCI key muscles, adjusted using the Dunn’s multiple comparisons correction. The Mann-Whitney U test was used to compare differences between two independent groups. Multiple Mann-Whitney tests were used to compare ranks and multiple comparison adjustments were performed using the false discovery rate and the two-stage step-up method of Benjamini, Krieger, and Yekutieli.^50, 51^

### Prediction of strength recovery after cervical SCI: the role of baseline MMS

To assess the predictive value of baseline MMS, we followed recent recommendations from studies on proportional recovery after stroke and performed descriptive statistics of strength recovery data^47^, implemented machine learning approaches^44^, controlled for ceiling effects^43^, and performed non-linear regression models using decision trees.^43, 52^ Random forest regressors were conducted using 50% of the dataset for training and 50% for testing with 100 trees (estimators). The random forest algorithm fitted several classifying decision trees on various sub-samples of the dataset and used averaging to improve the predictive accuracy and control over-fitting (in contrast to the original method).^53^ The non-linear regression model fit was assessed by the R^2^ and prediction error [average of abs(predicted-true)] and qualitatively by visual inspection of the regression lines.

### Prediction of strength recovery after cervical SCI: the role of additional muscle-specific features

Also following recent recommendations^43, 44^, we explored supervised machine learning models using random forest classifiers with additional baseline predictors – including quantitative multimodal electrophysiological assessment. The classifier was trained to predict motor recovery based on the change of MMS score between baseline and 48 weeks post-SCI (‘Recovery’: an increase of at least 1; ‘No recovery’: no change or decline). The following baseline features were used as predictors in the models (included features varied across models, as specified in relevant portions of the results): AIS, MMS, DST, LT, PP, Muscle identity, MEP amplitude, MEP score, SSEP amplitude, SSEP score, CMAP amplitude, F-wave persistence, and NCS score. Random forest classifiers were constructed using 100 trees (estimators). Evaluation was carried out using leave-one-muscle-out cross-validation. The performance of the random forest classifiers was assessed by the precision, recall/sensitivity, specificity, and F1-score of the predictions. Additionally, receiver operating characteristic and precision-recall curves^54^ were used to obtain the area under the curve (ROC AUC and PR AUC, respectively) - indicating the overall performance of the models as additional elements are added. The differences between the models including the electrophysiological features and models based only on clinical features were assessed using the McNemar’s test. To understand the importance of each feature to the prediction, we also reported the feature importance calculated as the decrease in node impurity weighted by the probability of reaching that node (computed using 50% of the dataset for training and 50% for testing; Supplementary Table 3). The node probability was calculated by the number of samples that reach the node, divided by the total number of samples. The higher the feature importance value the more important the feature.

### Role of CST and LMN integrity in impairment and recovery after SCI

Spearman correlation was used to explore the relationship between the change in MMS with change in MEP and CMAP amplitude. The relation between impairment and recovery of the spinal cord function was assessed using the motor component of the ISNCSCI, MEP amplitudes, and linear regression models. The ISNCSCI is a Likert-like scale, and thus is a summary of multiple Likert-like items, comprising ordinal data.^55^ We considered that the combination of multiple items renders the parametric statistical approaches applied here feasible.^43^ Mathematical coupling is an important statistical consideration when regressing the initially preserved motor functions against change scores (change in MMS from baseline to endpoint), which was extensively debated over the past years in the stroke recovery field.^43–45, 47^ Clustering algorithms commonly used in proportional recovery studies can bias the regression toward high values because of the low variability of the clustered data at the endpoint (after mathematically removing non-fitters).^43–45^ To address these issues, we refrain from using mathematical clustering, instead, we clustered the dataset based on physiological biomarkers^28, 29, 41, 46, 48^, i.e., the severity of SCI (AIS classification)^31^ or the presence(MEP^+^)/absence(MEP^-^) of MEP^28^.

### Data availability

Data is available upon reasonable request to EMSCI.

## Results

The clinical records of 748 research participants were reviewed in this study (26/748 with non-traumatic SCI). There were 599 males and 149 females, 261 classified as AIS A, 84 as AIS B, 155 as AIS C, and 241 as AIS D.^31^ Neurological level of injury ranged from C1 to C8 and the mean age was 46.5 years. Very acute clinical assessments were available for 440 participants and multimodal electrophysiological assessments were conducted in 203 (MEPs), 313 (SSEPs), and 280 (NCS) participants at the 4-week baseline (Supplementary Table 1).

### Segmental strength recovery in upper limb muscles

In accordance with previous findings^56^, individuals with AIS A and AIS B lesions have similar median UEMS early after the lesion (1w-4w) but participants with an AIS B regain more UEMS with time, 7 points at12w (P = 0.003, U = 2823), 9 points at 24w (P = 0.004, U = 2865) and 12 points at 48w (P = 0.001, U = 2726)). Individuals with an AIS B and C display similar upper limb recovery profiles (P > 0.008), but AIS C show greater UEMS at all time points, compared to AIS A (1w: 7 points, P = 0.002, U = 5788; 4w: 9 points, P < 0.001, U = 5013; 12w: 12.5 points, P = 0.001, U = 4178; 24w: 18 points, P < 0.001, U = 3962; 48w: 19 points, P < 0.001, U = 2847). Participants with an AIS D display the most upper limb strength recovery (P < 0.05; Figure 2A, B provide patterns of absolute UEMS scores and statistical comparison results, respectively).

**Figure 2.**
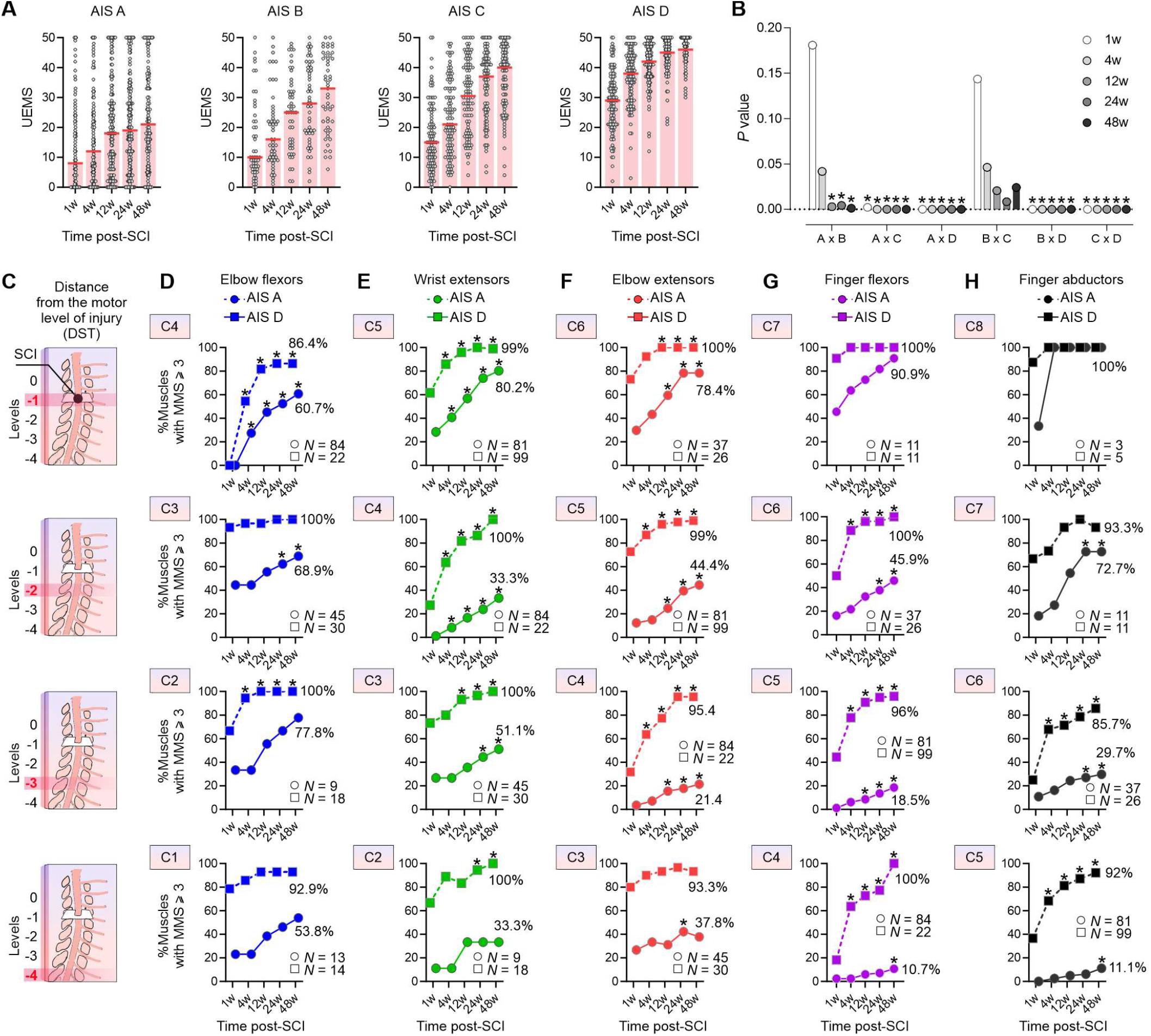
Strength recovery in upper limb muscles after cervical SCI. (**A**) The median UEMS in AIS A-D indicates some recovery independent of the SCI severity. While UEMS at onset are similar in AIS A and AIS B at 1w- 4w post-SCI the extent of recovery at 12w-48w is higher in AIS B. AIS C and D show different UEMS at onset and over time. (**B**) Statistical comparisons for the UEMS data in (A). (**C**) The distance between the motor level and the myotome (DST) was controlled in panels D-H. (**D-H**) In individuals classified as AIS A, the probability of the proximal muscles (i.e. elbow flexors, wrist extensors, and elbow extensors) achieve against gravity strength (MMS ≥ 3) was greater compared to hand muscles (i.e., finger flexors and abductors) – especially if the hand muscles are distant from the SCI (i.e., levels -3 and -4). Hand muscles also took longer to regain strength in individuals classified as AIS A. In participants classified as AIS D, the overall probability of upper limb muscles in reaching MMS ≥ 3 was greater (≈ 97%) compared to AIS A (≈ 50.6%), with the lowest probabilities for hand muscles . Multiple Mann-Whitney tests with multiple comparison adjustments using false discovery rate in **A, B**; *P < 0.05, McNemar’s tests in **D-H**. SCI = Spinal Cord Injury; AIS = American Spinal Cord Injury Association Impairment Scale; MMS = Muscle Motor Score; UEMS = Upper Extremity Motor Score. The insets in D-H describe the motor level of injury (C1-C8).

At the segmental muscle level, after controlling for the distance from the motor level (DST; Figure 2C), proximal muscles such as the elbow flexors and extensors show superior recovery compared to distal muscles such as the intrinsic hand muscles, if distant from the lesion. For example, at a DST of -3, the elbow flexors recover to a grade of 3 or more in 77.8% of individuals classified as AIS A and 100% of individuals classified as AIS D (Figure 2D); the wrist extensors recover to a grade of 3 or more in 51.1% of individuals classified as AIS A and 100% of individuals classified as AIS D (Figure 2E); the elbow extensors recover to a grade of 3 or more in 21.4% of individuals classified as AIS A and 95.4% of individuals classified as AIS D (Figure 2F); the finger flexors recover to a grade of 3 or more in 18.5% of individuals classified as AIS A and 96% of individuals classified as AIS D (Figure 2G); the finger abductors recover to a grade of 3 or more in 29.7% of individuals classified as AIS A and 85.7% of individuals classified as AIS D (Figure 2H). Overall, muscles from individuals classified as AIS A also take longer to recover strength to a grade 3 or more (P < 0.05). Although some statistical comparisons are hampered due to the low sample size of the very acute dataset (especially for AIS B/C – Supplementary Figure 1), additional analysis using the complete dataset (not including the 1w timepoint) corroborate these findings (Supplementary Figures 2-5). The bulk of the results indicates greater impact of the SCI and lesser strength recovery of distal muscles (finger flexors and abductors) compared to proximal upper limb muscles (elbow flexors), especially in individuals with sensorimotor or motor complete SCI, even after controlling for the distance from the lesion.

### Prediction of strength recovery after cervical SCI: the role of baseline MMS

Considering all AIS grades and all upper limb muscles, the prediction of strength recovery by baseline MMS is poor (R^2^ = 0.148; Figure 3A). On average, muscle-level analysis indicates that strength recovery (change in MMS scores from 4 to 48 weeks) is to some extent proportional if the initial baseline MMS is high (3-4), and plateaus between 1 and 2 points for lower baseline MMSs (Figure 3A). For individuals classified as motor complete (AIS A and B), strength recovery on average in the group of analyzed muscles is constant or inversely proportional if the baseline MMS is low – especially for hand muscles (Figure 3B, C). Proportional strength recovery is apparent on average in individuals classified as AIS C for elbow flexors, wrist extensors, elbow extensors, and finger flexors. Proportional recovery is also apparent for finger abductors if the initial impairment is low to mild (MMS = 2-4) but plateaus at 1.5 points if the baseline MMS is low (0-1; Figure 3D). Proportional strength recovery is evident for all muscles in individuals classified as AIS D (Figure 3E). Considering the different muscles and AIS grades, the non-linear regression using random forest algorithms using only the baseline MMS indicates good prediction of strength recovery for all muscles of AIS D participants with high R^2^ values and a prediction error of ≈ 0.5 points. Although the prediction is fair to good for some of the proximal muscles in individuals classified as AIS A/B/C, predicting strength recovery solely based on the initial motor impairment (baseline MMS) is overall poor, especially for distal muscles (R^2^ ≈ 0.1) (Figure 3F).

**Figure 3.**
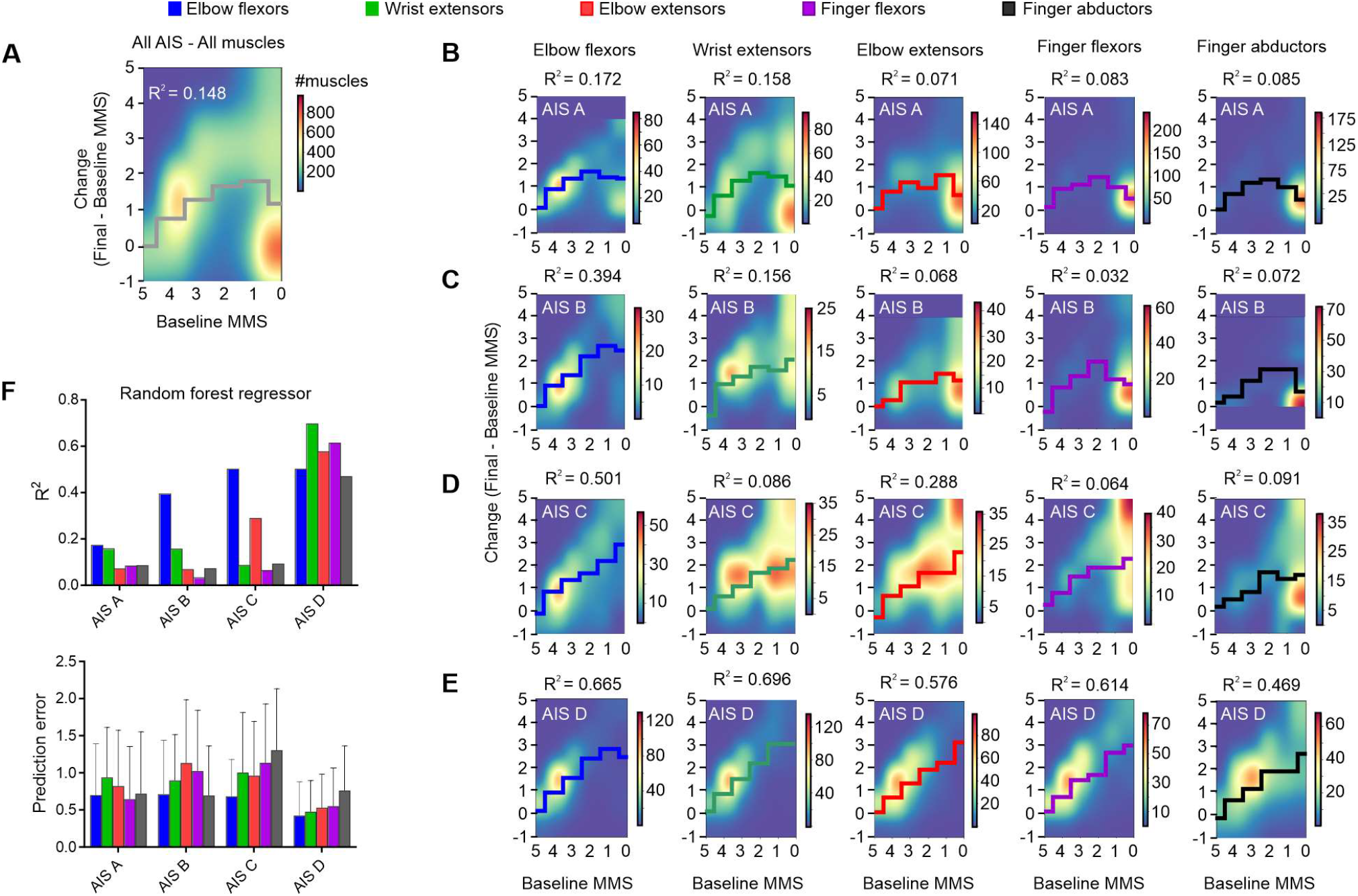
Prediction of strength recovery after cervical SCI: the role of baseline MMS. Baseline MMS is a good predictor of strength recovery at 1-year post-SCI for individuals with AIS D (high R^2^ values) but is in most cases a poor predictor for those with an AIS A/B/C. (**A**) Considering all AIS and muscles, the prediction of strength recovery using baseline MMS is poor (R^2^ = 0.148). (**B, C**) Individuals classified as AIS A or B show some degree of proportional recovery of upper limb muscles if the initial impairment is low, but strength recovery is constant or is inversely proportional if the initial impairment is high (especially for distal hand muscles). (**D**) In individuals classified as AIS C, proportional strength recovery is apparent for elbow flexors, wrist extensors, elbow extensors, and finger flexors. Proportional recovery is also evident for finger abductors if the initial impairment is low to mild (baseline MMS from 3-5) but is constant if the initial impairment is high (baseline MMS from 0-1). (**E**) Proportional strength recovery is evident for all muscles in individuals classified as AIS D. (**F**) Summary of the non-linear regression using random forest regressors indicates good prediction of strength recovery for all muscles of AIS D participants with a prediction error of ≈ 0.5 points. Although the prediction is fair to good for some of the proximal muscles in individuals with an AIS A/B/C, predicting late strength recovery solely based on the initial motor impairment is poor for distal hand muscles (R^2^ ≈ 0.1). Data is Mean + SD in **F**, bottom panel. SCI = Spinal Cord Injury; AIS = American Spinal Cord Injury Association Impairment Scale; MMS = Muscle Motor Score. Random forest regressor using 50% of the dataset for training and 50% for testing with 100 trees (estimators).

### Prediction of strength recovery after cervical SCI: the role of additional muscle-specific features

Given the inability to predict strength recovery using solely the baseline MMS in individuals classified as AIS A/B/C, especially in distal muscles, we explored additional segment-specific variables available in the ISNCSCI (i.e., LT, PP, and DST). A four-step approach using supervised machine learning models (Figure 4A and 5A) is employed to predict the presence or absence of recovery at the muscle level. First, we corroborate the importance of baseline AIS classification and MMS as predictive factors for strength recovery in SCI but expand it to the predictions of segmental strength recovery (Figure 4B; see Supplementary Table 3 for feature importance in each model).

**Figure 4.**
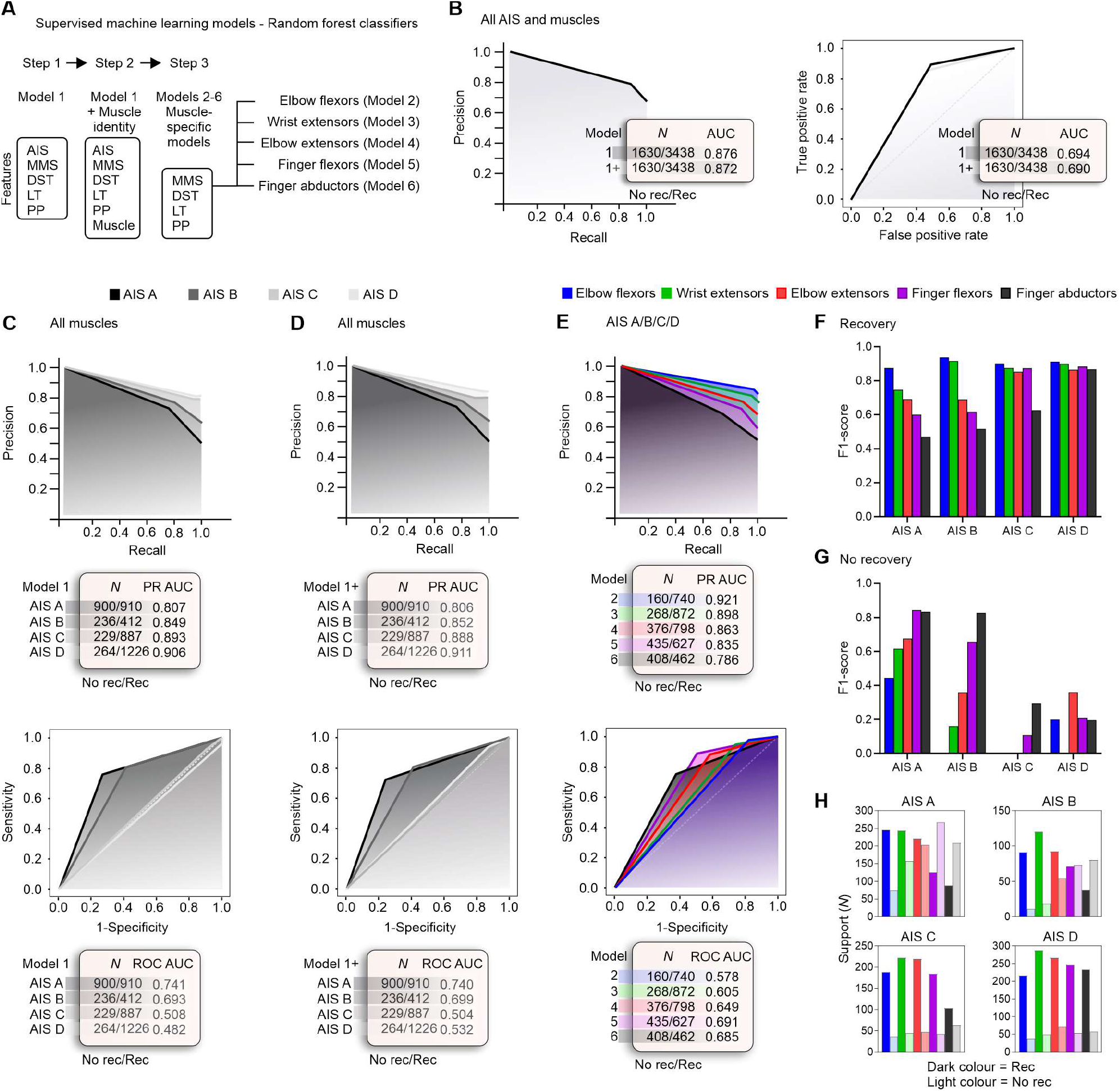
Prediction of strength recovery after cervical SCI: the role of additional muscle-specific features. The prediction of strength recovery displays a proximal-to-distal gradient in individuals with a sensorimotor complete lesion, where the strength recovery of distal hand muscles is hard to predict. (**A**) Supervised machine learning models: a three steps approach is utilized to understand the predictive factors for segmental strength recovery after cervical SCI. (**B**) We corroborate the importance of AIS and MMS in predicting recovery after SCI with a PR AUC of ≈ 0.87 and ROC AUC ≈ 0.69 (see Supplementary Table 3 for feature importance in each model). (**C**) AIS-specific models indicate it is harder to predict strength recovery in AIS A/B, compared to AIS C/D. (**D**) The addition of muscle identity as a feature does not increase the prediction performance of the AIS-specific models. (**E**) Muscle-specific models indicate a proximal to distal gradient, where the strength recovery of distal hand muscles is harder to predict compared to the proximal muscles. (**F**) In individuals classified as AIS A, the prediction of strength recovery displays a good performance for elbow flexors, a moderate performance is evident for wrist and elbow extensors, but the prediction of strength recovery is poor for hand muscles. The prediction of strength recovery is good for elbow flexors and wrist extensors but moderate for elbow extensors and poor for the hand muscles in participants classified as AIS B. In individuals classified as AIS C, the prediction is good for all muscles, except for finger abductors. Prediction of strength recovery in participants classified as AIS D shows good performance for all muscles. (**G, H**) Note that the muscle-specific models must be interpreted with caution because of the imbalanced datasets. AIS C/D and proximal muscles are trained with a predominance of muscles from the positive class (‘Recovery’ class), thus, perform poorly in classifying the negative class (‘No recovery’ class). Note: Random forest classifier using leave-one-muscle-out cross-validation. AIS = American Spinal Cord Injury Association Impairment Scale; AUC = Area Under the Curve; MMS = Muscle Motor Score; DST = Distance from the motor level of injury; LT = Light Touch sensation; PP = Pin Prick sensation; PR = Precision-Recall; ROC = Receiver Operating Characteristic.

The classification performance assessed by the PR AUC is overall higher for AIS C/D compared to AIS A/B, and the addition of the muscle identity as a feature does not afford an increase in the classification accuracy (All AIS: P = 0.095; AIS A: P = 0.944; AIS B: P = 0.832; AIS D: P = 0.924) or decreases the classification accuracy (AIS C: P = 0.003) (Figure 4C, D – upper panels). Note that the models for AIS C/D are imbalanced and have limited support for the ‘No Recovery’ class, leading to a high false positive rate and low performance on the ROC AUC (Figure 4C, D – lower panels). When constructing muscle-specific models, the prediction of strength recovery in elbow flexors displays a good performance on the PR AUC; a moderate performance is evident for wrist extensors and elbow extensors, but the prediction of strength recovery is lower for hand muscles (Figure 4E). Similar to the AIS models described in Figure 4C, D, the imbalance in support for the ‘No Recovery’ class of proximal muscles leads to a high false positive rate and low performance on the ROC AUC (Figure 4E – lower panel). Besides the limitations of the muscle-specific models (imbalance), it is evident that AIS A and B display a proximal to distal gradient, where the strength recovery of hand muscles is hard to predict. In individuals classified as AIS C, the prediction of strength recovery is good for all muscles except the finger abductors. Prediction of strength recovery in participants classified as AIS D shows good performance for all muscles (Figure 4F). Note that because of the imbalanced datasets, the muscle-specific models for AIS C/D were trained on a reduced number of muscles from the negative ‘No recovery’ class and perform poorly in predicting it (Figures 4G, H). Overall, the prediction of strength recovery displays a proximal-to-distal gradient in individuals with a sensorimotor complete lesion, where the strength recovery of distal muscles is hard to predict despite the inclusion of additional predictive variables in the model (i.e., muscle identity, LT and PP sensation). Although the residual strength at baseline (baseline MMS) is an important feature in predicting strength recovery especially in distal muscles (data not shown), it is not sufficient to afford a good prediction of strength recovery for distal muscles in individuals classified as AIS A/B/C (Figure 4F).

### Electrophysiological multimodal assessments improve the prediction of strength recovery in hand muscles after cervical SCI

Next, we sought to understand the predictive value of electrophysiological multimodal assessments in improving outcome prediction of strength recovery of distal muscles in individuals classified as AIS A/B/C (Figure 5A, B). Overall, biomarkers of CST and LMN integrity and somatosensory integration are increased at baseline in muscles that showed strength recovery 48 weeks after SCI (Table 1). Baseline MEP amplitudes of hand muscles with strength recovery are greater (P < 0.0001) and those hand muscles tend to have higher MEP scores (which indicates both high amplitude and low latency), compared to muscles with absent recovery (P < 0.0001). The SSEP and F-wave persistence of ‘Recovery’ muscles are also greater, compared to muscles with absent strength recovery 48 weeks after SCI (P < 0.0001 and P = 0.0009, respectively). This indicates the spinal cord is more responsive in integrating and transmitting neural input early after the injury in muscles regaining strength 1 year after SCI. CMAP amplitude was similar between ‘Recovery’ and ‘No recovery’ muscle groups, indicating the absence of LMN lesion in spinal segments innervating the abductor digiti minimi muscle (P = 0.316). Although SSEP and MEP assessments improve the prediction of strength recovery in hand muscles (Figure 5C-E), only the MEP increased the performance of the random forest classifier significantly (P = 0.015; Figure 5F).

**Figure 5.**
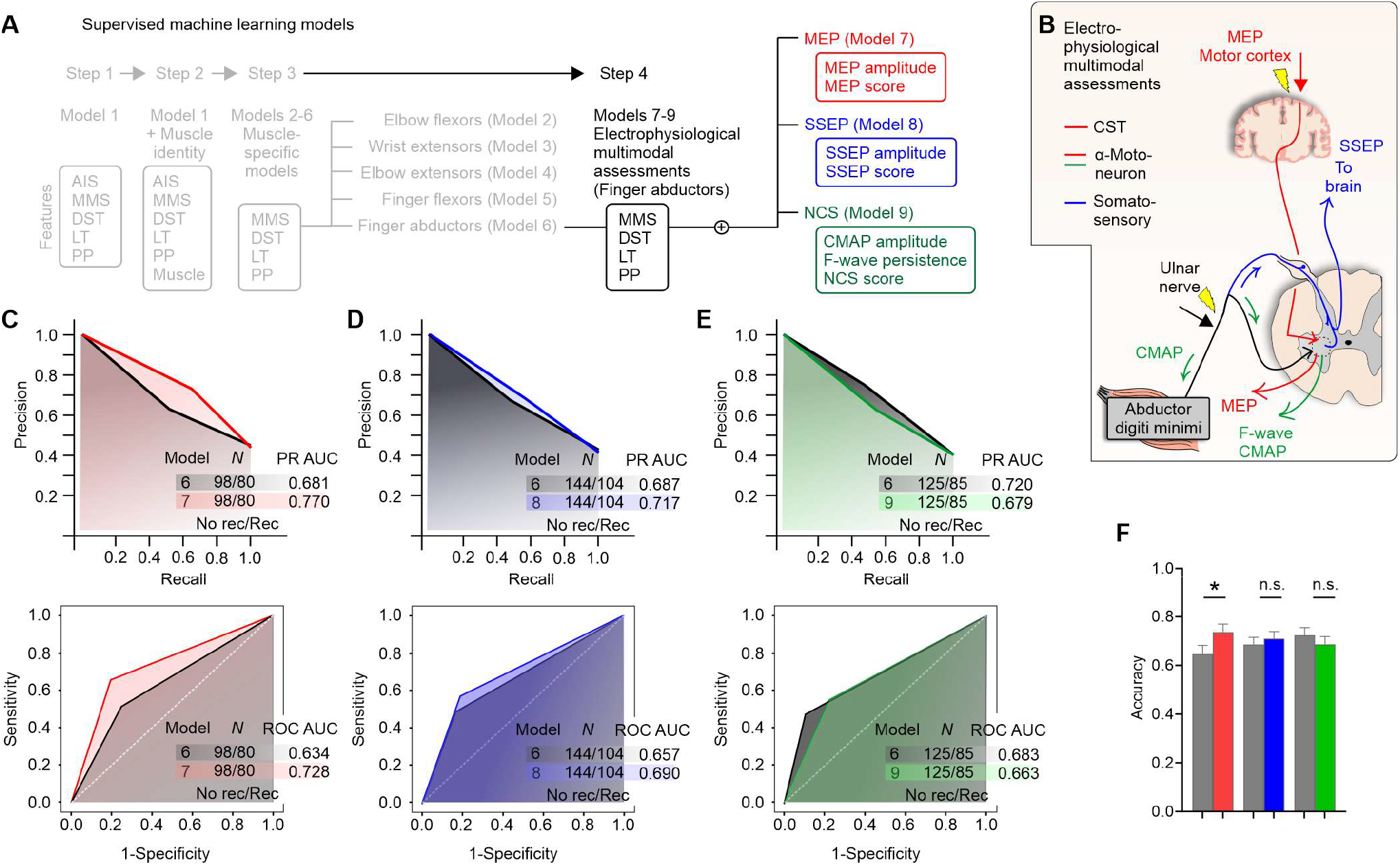
Prediction of strength recovery after cervical SCI: measures of spinal cord function (CST and LMN) integrity increase the classification performance of strength recovery of distal hand muscles in individuals with an AIS A/B/C. (**A**) Supervised machine learning models: a fourth step is utilized to understand the predictive factors for strength recovery in finger abductor muscles in AIS A/B/C. (**B**) Electrophysiological multimodal assessments of MEP, SSEP, and NCS of the distal muscles of the upper limb (finger abductors: abductor digiti minimi). MEP amplitude and latency at the abductor digit minimi muscle was used to quantify the CST and LMN integrity (red line). SSEP was measured over the scalp after stimulation of the ulnar nerve using needle electrodes (blue lines). Ulnar nerve stimulation was also used during the NCS to measure F-waves and CMAP at the abductor digit minimi (green line). (**C-E**) PR and ROC curves indicate that the overall classification performance assessed by the AUC is increased for the MEP and SSEP subgroups. The most important electrophysiological features are MEP amplitude, SSEP amplitude, and CMAP amplitude (data not shown). (**F**) Only the addition of the MEP features afforded a significant increase in the accuracy of the classification. Note: AIS = American Spinal Cord Injury Association Impairment Scale; AUC = Area Under the Curve; MMS = Muscle Motor Score; LT = Light Touch sensation; MEP = Motor Evoked Potential; CST = Corticospinal Tract; TMS = Transcranial Magnetic Stimulation; sEMG = surface Electromyography; DST = Distance from the motor level of injury; SSEP = Somatosensory Evoked Potential; EEG = Electroencephalography; LMN = Lower Motor Neuron; NCS = Nerve Conduction Studies; CMAP = Compound Muscle Action Potential; PR = Precision-Recall; ROC = Receiver Operating Characteristic. Random forest classifier using leave-one-muscle-out cross-validation. *P < 0.05, McNemar’s test (in F).

**Table 1.**
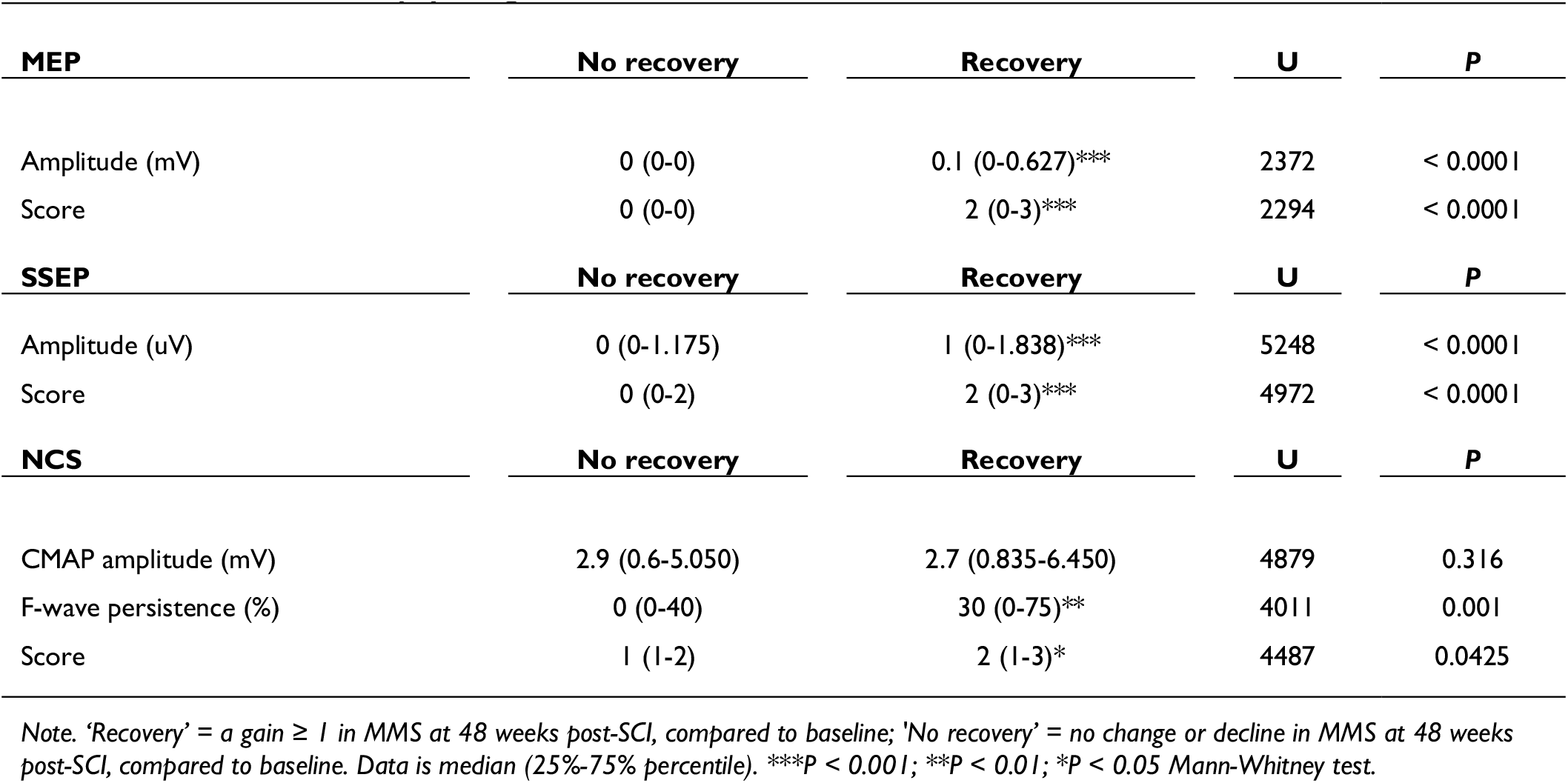
Outcome of electrophysiological examinations at baseline.

The strength recovery of hand muscles is limited in individuals classified as AIS A/B/C with an MEP^-^ at baseline, while the absence of MEP at baseline was indicative of greater impairment (MMS = 0) and reduced strength recovery in the finger abductors (Figure 6A). Finger abductor muscles with an MEP^+^ at baseline display greater variability in the initial motor impairment and strength recovery 48 weeks post-SCI, compared to MEP^-^ muscles (Figure 6B). Hand muscles with an MEP^+^ at baseline but with low MEP amplitude are less likely to regain strength (P < 0.0001; Figure 6C). Changes in muscle strength are accompanied by gains in MEP amplitude throughout the natural recovery process (P < 0.0001; Figure 6D). Although changes in hand muscles strength are also accompanied by an increase of CMAP throughout the natural recovery process, there is a weak association between CMAP at baseline and strength recovery 1-year after SCI (Figure 6E-K).

**Figure 6.**
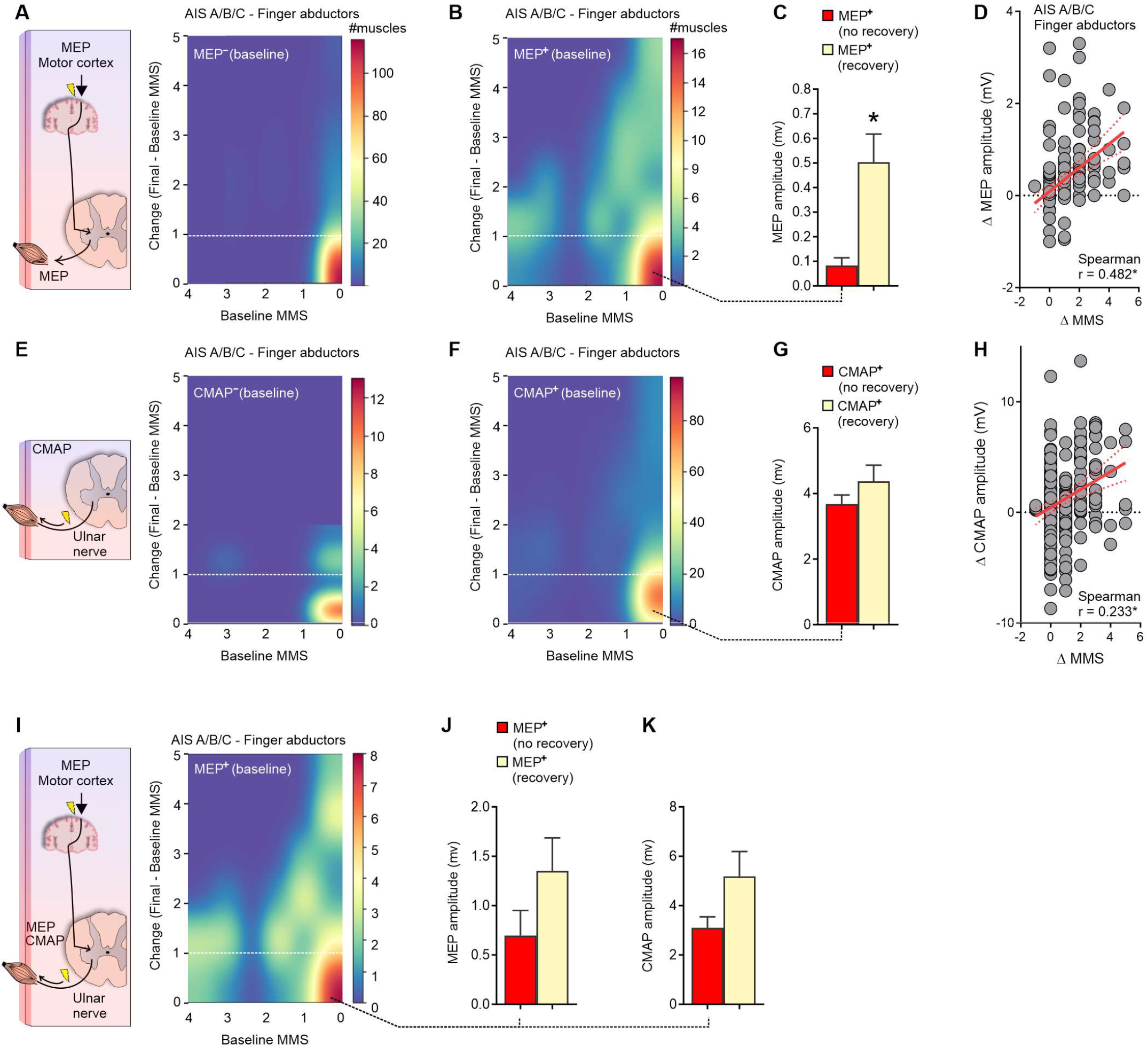
Corticospinal tract (CST) and LMN integrity in the *abductor digiti minimi* muscle. (**A**) In AIS A/B/C, finger abductor muscles with an MEP^-^ display limited motor recovery 1-year after SCI compared to (**B**) muscles with the presence of an MEP (MEP^+^) (yellow, above the hashed white line), including muscles with absent MMS at baseline (rightmost part of the heatmap). (**C**) The binary classification of MEP^+^ muscles that shows strength recovery or not was used to cluster the baseline MEP amplitude. This analysis indicates that higher MEP amplitudes were associated with increased strength recovery. (**D**) The strength recovery of finger abductors (Δ MMS) is accompanied by changes in MEP amplitude (Δ MEP amplitude) throughout the natural recovery process. (**E-H**) Conversely to MEP, the presence of CMAP at baseline is not strongly associated with motor recovery of the finger abductor muscles 1-year post-SCI in AIS A/B/C. (**I-K**) In a subgroup of individuals where both MEP and NCS studies were conducted, it is evident that muscles with an MEP^+^ at baseline and with strength recovery 1 year after SCI also show greater CMAP intensities at baseline. Note: MMS = Muscle Motor Score; AIS = American Spinal Cord Injury Association Impairment Scale; CMAP = Compound Muscle Action Potential; MEP = Motor Evoked Potential. Data are shown as Mean ± SEM in **C,G,J,K** to improve visualization. Outliers were left out of **D** (5 data points), **H** (2 data points) to improve visualization. *P < 0.05, Spearman correlation (in **D,H**).

### CST and LMN integrity indicates impairment and recovery after SCI

Damage to the spinal cord results in muscle weakness below the lesion, with pronounced effects on hand muscles. The residual spinal cord function may be measured by the residual strength of muscles or the CST and LMN integrity (assessed by the MEP) (Figure 7A). Individuals with absent MEP at baseline (MEP^-^) lack CST or LMN integrity and display greater initial impairment with limited strength recovery 1-year post-SCI, as measured by the total motor score (P = 0.492). Baseline CST and LMN integrity (MEP^+^) supported motor recovery at variable degrees (P < 0.0001; Figure 7B). Most individuals with MEPs^-^ were classified as AIS A at baseline, on the other hand, as AIS D if an MEP^+^ was evident at baseline (Figure 7C). Indeed, individuals classified as AIS B/C/D show recovery proportional to the largest possible improvement (AIS B: P = 0.016; AIS C: P = 0.031; AIS D: P < 0.0001; Figure 7D). No relationship between the largest possible and actual improvement is evident for individuals with a sensorimotor complete lesion (AIS A: P = 0.662). This analysis indicated that individuals with an MEP^-^ do not show proportional recovery to the largest possible improvement and are predominantly classified as AIS A (44.4%). The bulk of these results suggests that strength recovery can be predicted using solely baseline total motor score in AIS D and reinforces the importance of MEP in predicting the recovery of spinal cord function in AIS A/B/C.

**Figure 7.**
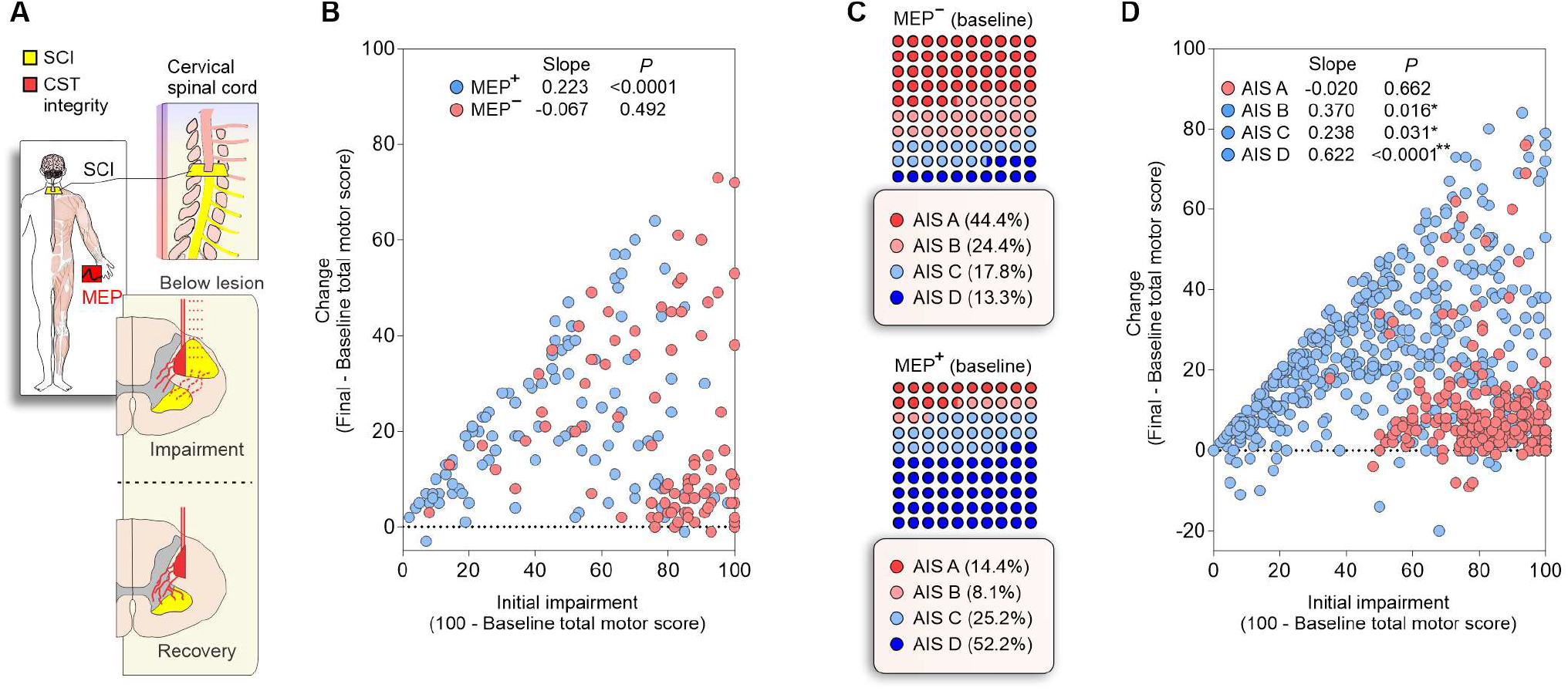
CST and LMN integrity (as assessed by MEP) indicates impairment and recovery after SCI: individuals classified as AIS A and impaired spinal cord function (CST and LMN integrity) display limited motor recovery. (**A**) Cervical SCI (yellow) may damage spinal cord structures and affect the spinal cord functionality below the level of injury with respective weakness of the innervated muscles. Volitional strength and strength recovery is dependent on the residual spinal cord function (red), here quantified by the residual muscle strength and MEP amplitude. (**B**) Individuals with absent MEP (MEP^-^) display greater damage to the descending pathway (evidenced by the greater initial impairment) and limited recovery of motor function of the spinal cord (P = 0.492). The presence of an MEP (MEP^+^) indicates variable levels of spinal cord or LMN damage and recovery (P < 0.0001). (**C**) Individuals with an MEP^-^ at baseline were predominantly classified as AIS A, and individuals with an MEP^+^ predominantly classified as AIS D. (**D**) Strength recovery from baseline (4 weeks) to 48 weeks after SCI is shown as change in the total motor score of the ISNCSCI. For individuals classified as AIS B/C/D (blue circles), recovery is proportional to the available improvement. In AIS D, the regression represents the relationship between available (x) and actual (y) improvement (y=0·62x). No relationship exists between available and actual improvement for sensorimotor complete lesions (AIS A; red circles). Note: AIS = American Spinal Cord Injury Association Impairment Scale; MEP = Motor Evoked Potential; LMN = Lower Motor Neuron.

## Discussion

Natural recovery after cervical SCI relates to the segmental innervation and follows a proximal-to-distal gradient in which distal muscles of the upper limb show limited and delayed strength recovery compared to proximal muscles. In addition, the recovery of hand muscle strength depends on the severity of spinal cord damage. In more affected individuals, non-linear interactions between residual muscle strength and the amount of recovery 1-year post-SCI challenge predictions while on average recovery is proportional to the impairment if the initial impairment is low to mild (baseline MMS from 3-4). Residual baseline strength was insufficient to predict motor recovery in severely impaired individuals – especially in the hand muscles, even when additional clinical features were included in the models. Baseline electrophysiological assessments soon after the SCI provide measures of CST and LMN integrity, and increased the prediction performance in the abductor digiti minimi. In the hand muscles, stronger MEPs at baseline were positive indicators of recovery, and positive changes in MEP amplitude were associated with strength recovery over time. Overall, our data support the importance of CST and LMN integrity in indicating spinal cord dysfunction and recovery, here evidenced by the residual strength and MEP at baseline. Nonetheless, integrity of these descending pathways at baseline was not always related to a good motor recovery prognostic of the hand muscles. Some hand muscles with MEP^+^ at baseline did not show motor recovery 1 year after SCI. This indicates that other variables may explain the variance of the outcomes during the recovery process and neurorehabilitation needs to be optimized for those muscles with potential to recover early after SCI.^57^

Upper limb motor recovery after SCI has been extensively studied over the past decades but most of the studies did not focus on the segmental approach described here.^5,7, 11–13, 58–60^ Greater specificity in understanding the recovery of upper limb muscles is desirable for detecting subtle changes, because even small gains in upper limb function can have important repercussions on independence and quality of life.^8, 61^ Here, we expand the natural recovery to the muscle level and provide evidence of limited and delayed recovery of distal compared to proximal upper limb muscles. The greater impact on hand muscles after controlling for distance from the lesion may be explained by the amount of CST projections to spinal motoneurons, which are greater in distal muscles compared to proximal muscles.^20–22^ Our data also indicates a low prevalence of LMN damage in motoneuron pools innervating the distal hand muscles (10.5% of abductor digit minimi muscles in AIS A/B/C showed a CMAP amplitude of 0 mV at the 4-week timepoint), supporting the idea of UMN lesions primarily accounting for our results (Table 1). Nonetheless, the assessments available in our dataset did not provide a complete assessment of LMN function for all muscles, and therefore the inability to fully account for LMN damage in the predictive models is a limitation of our study.

We hypothesize the residual CST projections to distal muscles to be the major player in muscle-specific impairment and recovery of volitional movements. This is supported by the importance of residual muscle strength (AIS D) and MEP (AIS A/B/C) for predicting strength recovery of hand muscles. Interestingly, the strength recovery of hand muscles was also associated with stronger MEP amplitudes at baseline and throughout the natural recovery process. This increase in MEP amplitude with recovery may reflect extensive spontaneous plasticity of CST projections, previously evidenced in a primate model of SCI.^62^ Our findings are also supported by previous clinical findings indicating that greater MEPs at baseline are associated with increased recovery of the MEP during the first year after SCI.^23^ This plasticity of the residual CST projections may involve transsynaptic mechanisms rather than sprouting of spinal cord axons, which is not well observed in preclinical studies. Importantly, here, the addition of MEP as a feature increased the performance of the predictive models of strength recovery for the hand muscles. Thereby, given the high priority in regaining hand function in tetraplegia^2^, patients would benefit from additional electrophysiological assessments early after the SCI. Indeed, if this residual CST functionality goes unnoticed early after the injury, the opportunity to strengthen these projections may be lost^63, 64^ – likely explaining why some participants displayed positive signs of spinal cord function integrity at baseline but displayed absent recovery with time. Of note, we cannot fully account for the role of other descending spinal tracts, which have differential effects on proximal and distal upper limb control^65–67^. For example, in SCI, it is known that the reticulospinal tract assists hand control during gross finger manipulations.^68^ Likewise, motor unit plasticity at the muscle level may also play a role in MEP amplitude changes. These are limitations of our study and prevent definitive conclusions about the role of the CST in our results.

Notwithstanding the greater amount of CST projections to spinal motoneurons in distal muscles compared to proximal muscles^20–22^, the lack of proximal-to-distal gradient in the number of efferents that leave the spinal cord suggests that only a few motor units control fine hand movements.^33^ There are over 20 intrinsic muscles in the hand^69^ responsible for fine control of several degrees of freedom, which are innervated by only ≈ 1,700 motoneurons^33^ controlled by an immense neural network in primary motor areas of the cortex.^18, 32^ Therefore, it has been suggested that dexterous control over multiple degrees of freedom is not achieved by a finer recruitment of motor neurons in hand muscles compared to larger muscles with much grosser actions.^33^ Here, the pronounced effect of the SCI on hand muscles may reflect this reliance on CST projections, indeed evidenced by the importance of residual muscle strength and MEP in predicting strength recovery of intrinsic hand muscles.

The addition of SSEP did not significantly increase the performance of the prediction of strength recovery for the hand muscles. Given the predominance of sensory axons with respect to motor axons in the mixed peripheral nerves, and the increase of this ratio when moving from proximal to distal upper limb muscles^33^, it is reasonable to think of the sensory information as paramount to normal hand function. We suggest that the sensory component is less important to strength recovery than it is to function. The enhanced performance of models predicting function in SCI when incorporating SSEP^4^ and the reduced ability to control fine hand movements in the absence of touch and proprioceptive sensory input^70^ support this conclusion.

This is the first study exploring proportional recovery in SCI under the well-established framework developed for stroke.^28–30, 41–48^ Besides the obvious differences between these lesions, a common aspect is the importance of the damage to the CST. The lack of proportionality in strength recovery and prevalence of MEP^-^ in the hand muscles suggest that individuals classified as AIS A more commonly lack CST integrity (‘non-fitters’ to the proportional rule) - with greater upper limb motor impairments associated with non-linearities in the strength recovery profile. Proportionality was somewhat evident for the proximal muscles in people classified as AIS A/B/C, but the strength recovery of distal muscles was hard to predict. Indeed, hand muscles were the most important players in breaking the proportionality for those individuals. The break or inversion of proportionality when the residual strength is low, or an MEP is absent, reflects the importance of preserved CST projections for motor recovery after SCI. The similarities between the UEMS of AIS A and B at 1-4 weeks but the greater recovery of AIS B at 12-48 weeks after SCI also supports the importance of CST and LMN integrity for upper extremity motor recovery. The fact that individuals with AIS B show proportional recovery in their total motor score (y=0.37x, Fig. 7D) with a respective lower prevalence of MEP^-^ at baseline compared to AIS A (AIS A: 44.4%; AIS B: 24.4%) suggests a role for preserved CST projections and LMN integrity for upper extremity recovery even in AIS B. In individuals with AIS D, we suggest that the relationship between residual strength and strength recovery exists, and the prediction of motor recovery is possible using only the baseline MMS (y=0.62x). This variable upper limb recovery in AIS-based subgroups agrees with recent findings also indicating that different subgroups present distinct recovery profiles in stroke.^71^ Because it is thought that the proportional recovery from motor impairment reflects a ubiquitous neurobiological process, likely related to the biological limits of structural and functional plasticity^29^, these non-linearities may indicate the constraints for recovery. Indeed, given the importance of residual strength, CST projections, and LMN integrity for proportional strength recovery in SCI, we suggest these are the basis for the strength recovery that occurs at different proportionality slopes. Finally, the limited recovery in SCI compared to stroke may reflect the direct lesion to the CST in SCI, and the fact that in stroke, the redundancy of the sensorimotor circuitry within the brain allows compensation.^72, 73^

An important question that arises from this analysis is how to optimize the strength recovery of different muscles. The answer to this question is multifaceted but it is surely an important step to understand that different muscles may follow distinct recovery profiles and the predictive value of baseline assessments. The adjunct of electrophysiological measures of volitional activity (EMG) or CST and LMN integrity (MEP) is of utmost importance for weaker muscles – especially for those with absent MMS at baseline. In future studies, identifying the potential to recover strength may help to tailor rehabilitation to novel and intensive approaches, for example, anti-NOGO therapy^74^ to release the brakes of plasticity in the spinal cord and promote axonal sprouting, and paired associative stimulation^75^ - to induce long-term plasticity in the CST projections. The identification of muscles with the potential to recover early after the SCI will allow the administration of novel and promising therapies during the optimal time window for recovery. The enrollment in such rehabilitation programs must also be combined with intense rehabilitation to optimize recovery^76^ and avoid aberrant plasticity.^77^

There is a continuous effort in improving clinical trial design and outcomes in SCI.^78^ The most common method to quantifying the effects of treatment is by adding up several ordinal endpoints to form a single overall score (e.g. UEMS), but this may mislead associations and reduce statistical power.^79^ There is a compelling interest in statistical models specifically designed for the analysis of complex ordinal endpoints, such as autoregressive transitional ordinal models.^79^ Here, we employed machine learning methods using non-linear regressions and classification to account for the complexity of the ordinal outcomes. Future clinical trials should also consider the use of baseline-adjusted models, where the stratification based on baseline variables would improve the analysis of complex trial designs.^80^ Here we show how the baseline MMS and MEP are important in predicting recovery, especially for distal hand muscles. In a limitations section, we discuss the advantages, limitations, and future directions of the muscle-specific approach applied here.

Here, we investigated segmental strength recovery after cervical SCI. We demonstrated that recovery profiles of ISNCSCI key muscles are dependent on the distance from the lesion, baseline muscle strength, and SCI severity assessed by the AIS. Additionally, we show that lower recovery is present in the distal compared to proximal upper extremity muscles. While muscle strength recovery was proportional to the initial muscle strength in individuals classified as AIS D, the proportionality was lower for AIS B/C and broke for AIS A. Specifically the prediction of hand muscles strength benefited from the addition of MEP as a proxy of spinal cord function, i.e., CST and LMN integrity. To further determine the therapeutic consequences, the latter findings should be integrated into clinical assessment strategies following cervical SCI.

### Limitations, advantages, and future directions of research

Some limitations, advantages, and future directions of research are explored in this section.

#### Segmental distribution of motoneurons innervating upper limb muscles

The segmental distribution of motoneurons innervating upper limb muscles is one limitation when using the motor component of the ISNCSCI rating - which is not comparable between segments (as they are non-linear in intervals).^31^ For example, the α-motoneurons innervating the elbow flexors (biceps brachii) are located at several spinal cord segments (C5-C8), compared to hand muscles (constrained to C8-T1). Also, the motor deficits at different levels of the motor component of the ISNCSCI are not comparable and not necessarily related purely to voluntary motor control nor function.

#### Selection bias and limitations of the neurophysiological assessments

The calculated segmental percentages of recovery may differ due to selection bias, i.e., the prevalence of C4 and C5 lesions, and with the initial degree of segmental paralysis. Thus, it is more likely that proximal muscles are more affected by lower motor neuron (LMN) lesions, compared to hand muscles. The inability of our machine learning models in predicting proximal muscles that do not recover strength (Figure 4G) may be related to the presence of LMN lesions in muscles with absent recovery. Future studies should investigate if the assessment of other nerves, e.g., the musculocutaneous nerve, could improve the prediction of proximal muscles that do not regain strength after SCI. We also pinpoint limitations of anatomical nature, such as not accounting for the role of other spinal tracts.^65–68^ Given the complexity of the cervical spinal neuroanatomy it is important to consider the role of other spinal tracts, e.g., the concept of medial (bilateral, indirect/proprio- and reticulo-spinal) versus lateral (unilateral CST) motor systems in controlling proximal versus distal upper limb muscles, respectively.^65–67^ These spinal tracts are neuroanatomically different in terms of grey matter and somatotopy, indicating that projections contralateral/dorsolateral to distal limb α-motoneurons are less redundant than bilateral ventromedial spinal projections to proximal/trunk α-motoneurons. For example, in SCI, it is known that the reticulospinal tract assists hand control during gross finger manipulations.^68^ Another aspect to take into account is the number of α-motoneurons innervating upper limb muscles. It is known from non-human primate experiments that the number of α-motoneurons projecting to proximal upper limb muscles is greater (biceps brachii ≈ 1,020; triceps brachii ≈ 1,293; extensor carpi radialis and ulnaris ≈ 1,022) compared to distal hand muscles (intrinsic hand muscles: flexor pollicis and abductor pollicis brevis ≈ 122; lateral lumbricalis ≈ 68; first dorsal interossei ≈ 184).^81^ Thus, a lesion of similar size may have more pronounced effects on distal hand muscles, compared to proximal upper limb muscles. In this line of thought, this study contributes to a deeper understanding of the motor impairment and recovery at the segmental, not person, level. The use of the segmental analysis described here will circumvent current difficulties in characterizing the motor impairment at the person level (summed UEMS) by affording the description of the specific motor impairment at each muscle, e.g., muscles with flaccid paralysis (LMN lesion) or lacking control or with spasticity (UMN lesion).

#### Final considerations on “why it is so hard to regain strength and predict the strength recovery in distal hand muscles?”

The stronger monosynaptic (fast) response of distal forelimb muscles in relation to proximal upper limb muscles also supports the reliance of distal muscles on direct CST projections.^17, 82^ Around 1,600 motor axons innervate the elbow flexors (i.e. main branch of the musculocutaneous nerve), while a similar number of motor axons (about 1,700) are required for the higher number of muscles in all of the distal hand muscles (i.e., ulnar and median nerves at the arm level).^33^ This evidence suggests that a similar number of motor units innervate fewer muscles in the proximal arm muscles, compared to distal hand muscles. Thereby, proximal upper limb muscles display lesser direct CST projections^17^ and a greater proportion of motor axons per muscle^33^, which allows for more redundancy and compensation when the control of some motor units is lost after SCI. For example, the elbow flexors (C5) may lose some of the motor units but still be able to produce partial strength using the remaining motor units, which will expand the motor unit size to increase strength production with time.^83^ On the other hand, partial loss of function at the C8 level may drastically impair some distal hand muscles because each hand muscle is innervated by a small number of motor units, which strongly rely on the CST projections. Another evidence of redundancy in the innervation of proximal upper limb muscles is the above-mentioned contribution of several levels of the spinal cord to proximal upper limb muscles compared to distal hand muscles.^84^ Thus, in the distal hand muscles, there is no opportunity for expansion of the motor unit size at the muscle level, nor redundancy or alternative pathways at the spinal cord level – given the reliance on direct CST projections. Together, this evidence may explain why it is so hard to regain strength and predict the strength recovery in distal hand muscles (Supplementary Figure 1C, D).

#### Additional limitations

The following factors were not considered in the analysis.

1. other factors related to muscle anatomy and mechanics, such as pennation angle, fiber type composition, and cross-sectional area;
2. control for the rehabilitation provided to each individual during the natural recovery process - it is known that rehabilitative efforts initially focus on improving the proximal upper limb strength, which initially involves training anti-gravity muscles as the individuals learn how to transfer during the activities of daily living;
3. account for other anatomical systematic differences between subgroups, such as spinal syndromes (e.g., central cord or Brown-Sequard syndromes).

## Data Availability

All data produced in the present study are available upon reasonable request to the authors.

## Acknowledgments

We would like to thank all the research volunteers and the EMSCI study group.

## Supplementary Material

### Materials and methods used in the electrophysiological multimodal assessments of the hand muscles

#### Motor evoked potentials

TMS was used to quantify the CST and LMN integrity by the motor evoked potential (MEP) amplitude and latency measured using sEMG of the abductor digiti minimi muscle at both sides of the body. The following TMS stimulation parameters were used: (Coil) double cone if available, or else ring-shaped pancake coil; (Position) Double Cone was 45° inclined to the contralateral side over C3 or C4 respectively (M1 was identified by C3 and C4 of the 10/20 EEG system); (Position) Ring-shaped pancake coil placed over C3, inclined 30-45° to the contralateral side and placed over C4, 30-45° inclined to the contralateral side; (Background sEMG) isotonic contraction of the abductor digiti minimi muscle (20% of maximal contraction) where possible; (Intensity) threshold 1,5 fold. For recording the MEP at the abductor digiti minimi muscle: (Electrodes) surface electrodes; (Positioning) active electrodes were over muscle body of the abductor digiti minimi muscle and V metacarpophalangeal joint; (Ground electrode) between stimulation and recording; (Impedance) < 5kOhm; (Filters) bandpass between 10Hz-2000Hz; (Recording time) 100ms; (Reproductions) 3-5 clear reproductions (latency variations within 0.5 ms; amplitude variations within 20%). Data analysis included latency (stimulation-onset in ms of the fastest response) and amplitude (baseline to maximal negative peak in µV of the largest amplitude). The values ranged from 10-50ms (latency) and 0-20mV (Amplitude).

#### Somatosensory evoked potentials

EEG was used to quantify SSEP amplitude and latency after neuromuscular electrical stimulation (NMES) of the ulnar nerve at both sides of the body. The following stimulation parameters were used: (Pulse) square wave with a pulse width = 0.2ms (increased up to 0.5ms in cases of unsatisfying responses) and frequency = 3Hz; (Intensity) motor threshold; (Stimulation site) ulnar nerve at the ulnar side of the pulse. For recording: (Electrodes) disposable needle electrodes; (Positioning): C3 (for stimulation on right pulse) or C4 (for stimulation on the left pulse) against Fz; (Ground) in between stimulation and recording; (Impedance) < 5kOhm; (Filters) Cortical = bandpass between 10Hz-2000Hz and Peripheral = bandpass between 50Hz-2000Hz; (Recording time) 100ms; (Reproducibility) > 1 clear reproduction (difference between the two sets of averaged responses no bigger than 0.5ms for latency and 20% for amplitudes). Data analysis included latency (minimal N20, corresponding P30 and N9, all in ms) and amplitude (N20/P30 in µV). The latency values ranged from 15-50ms (N20), 20-50ms (P25) and 52-25ms (N9); and the amplitude values between 0-10µV.

#### Nerve conduction studies

NMES was also used to quantify compound muscle action potential (CMAP) amplitude, nerve conduction velocity (NCV), and F-wave persistence in nerve conduction studies (NCS) at both sides of the body. Similar to SSEP, the ulnar nerve was electrically stimulated. The following stimulation parameters were used: (Stimulation) pulse with = 0.1-0.2ms (increase to 0.5ms in case of unsatisfying response); (Intensity) supra maximal; (Stimulation site) ventral wrist, ulnar sides [Cathode distal] and proximal sulcus ulnaris [Cathode distal]. For recording: (Electrodes) surface electrodes; (Positioning) active electrodes were over muscle body of the abductor digiti minimi muscle and the V metacarpophalangeal joint; (Ground) between stimulation and recording; (Impedance) < 5kOhm; (Filters) band-pass between 5-10.000Hz. Data analysis included latency (distal motor latency: stimulation onset in ms), amplitude (CMAP amplitude: baseline to maximal negative Peak in mV) and NCV. The distal motor latency values ranged from 1.5-10ms, the CMAP amplitude from 0-50mV and the NCV from 20-100m/s. For the F-wave persistence analysis the same parameters were used except the filter (band-pass filter between 100-10.000Hz), 10-20 stimulations were conducted, and values ranged from 20-100ms (latency) and 0-100% (F-wave persistence). The values for F-wave persistence were a percentage of the total stimulations in which an F-wave could be elicited.

## Supplementary Figures and Tables

**Supplementary Figure 1.**
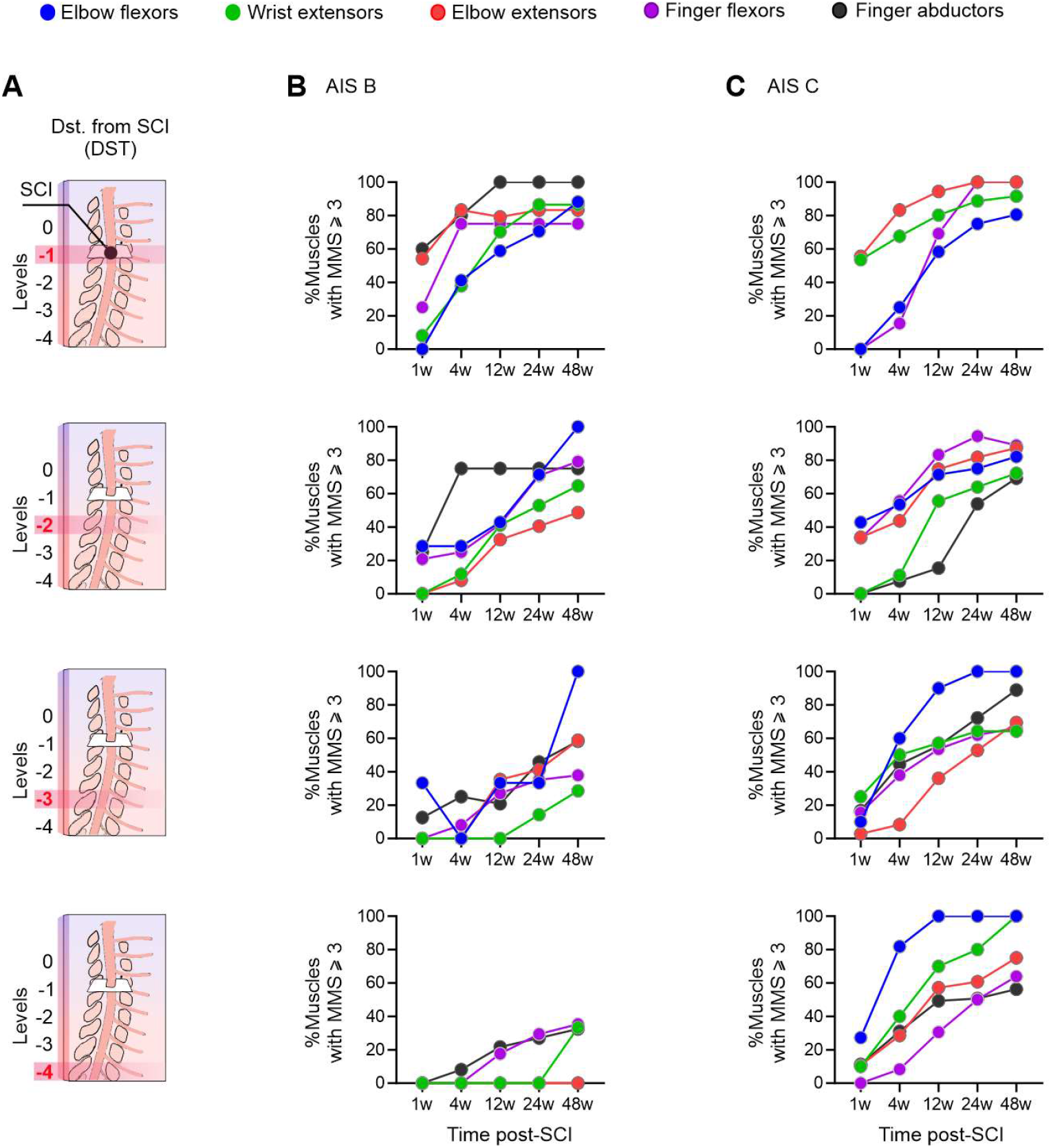
Strength recovery in upper limb muscles after cervical SCI. (**A**) The distance between the SCI and upper limb muscles (DST) was controlled in panels B, C. (**B, C**) In individuals classified as AIS B or C, the probability of the proximal muscles (i.e. elbow flexors, wrist extensors, elbow extensors) achieve against gravity strength (MS ≥ 3) was greater compared to distal muscles (i.e., finger flexors and abductors) – especially if the respective muscle is distant from the SCI (i.e., levels -3 and -4).

**Supplementary Figure 2.**
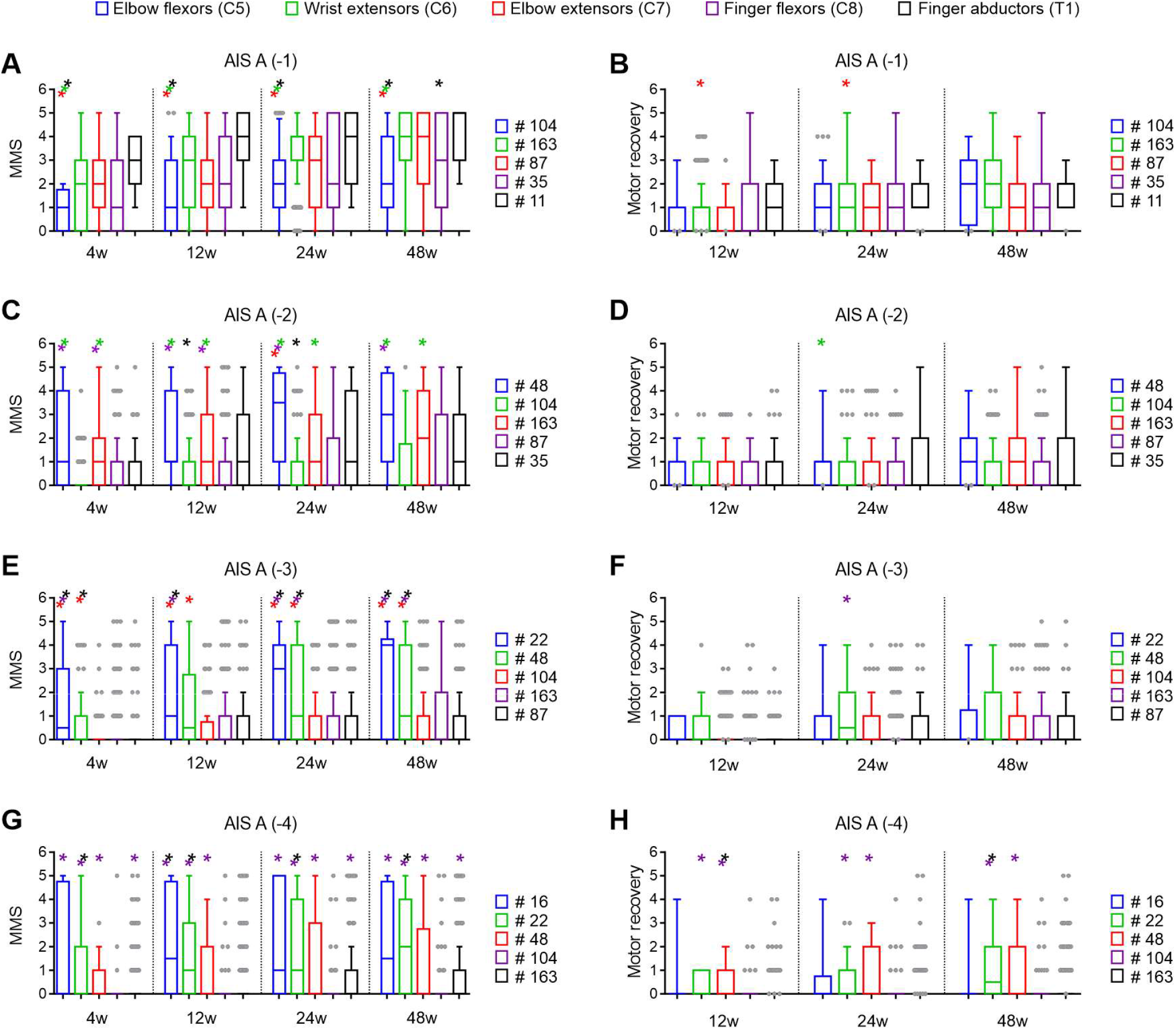
Motor recovery of muscles below the motor level of injury in AIS A individuals. (**A**) Muscles that are one level below the injury (−1) show distinct responses after the injury, the elbow flexors were more impaired. (**B**) All muscles at this distance from the injury recover to a similar extent 48 weeks post-SCI. (**C**) When muscles are two levels below the injury (−2) the SCI seems to display more effect on distal muscles such as finger flexors and abductors. (**D**) All muscles at this distance from the injury recover to a similar extent 48 weeks post-SCI. (**E, G**) The effect described in D becomes more apparent when the muscles are further below the level of injury (−3 or −4 levels), the lesion has more impact over distal hand muscles in comparison to proximal muscles. (**F, H**) Strength recovery of these muscles seems to occur more at wrist and elbow extensors compared to finger flexors and abductors (**H**). Kruskal-Wallis test with Dunn’s multiple comparisons test: *P < 0.05 relative to elbow flexors (*blue), wrist extensors (*green), elbow extensors (*red), finger flexors (*purple) or finger abductors (*black). Note: data are median (line in the middle of the box), the first quartile forms the bottom, and the third quartile forms the top of the box, whiskers represent the 5-95 percentiles; any data beyond the whiskers are shown as points (grey dots); the numbers on the right of each panel indicates the number of muscles available for each analysis. SCI = Spinal Cord Injury; AIS = American Spinal Cord Injury Association Impairment Scale; MMS = Muscle Motor Score; C = Cervical level; T = Thoracic level.

**Supplementary Figure 3.**
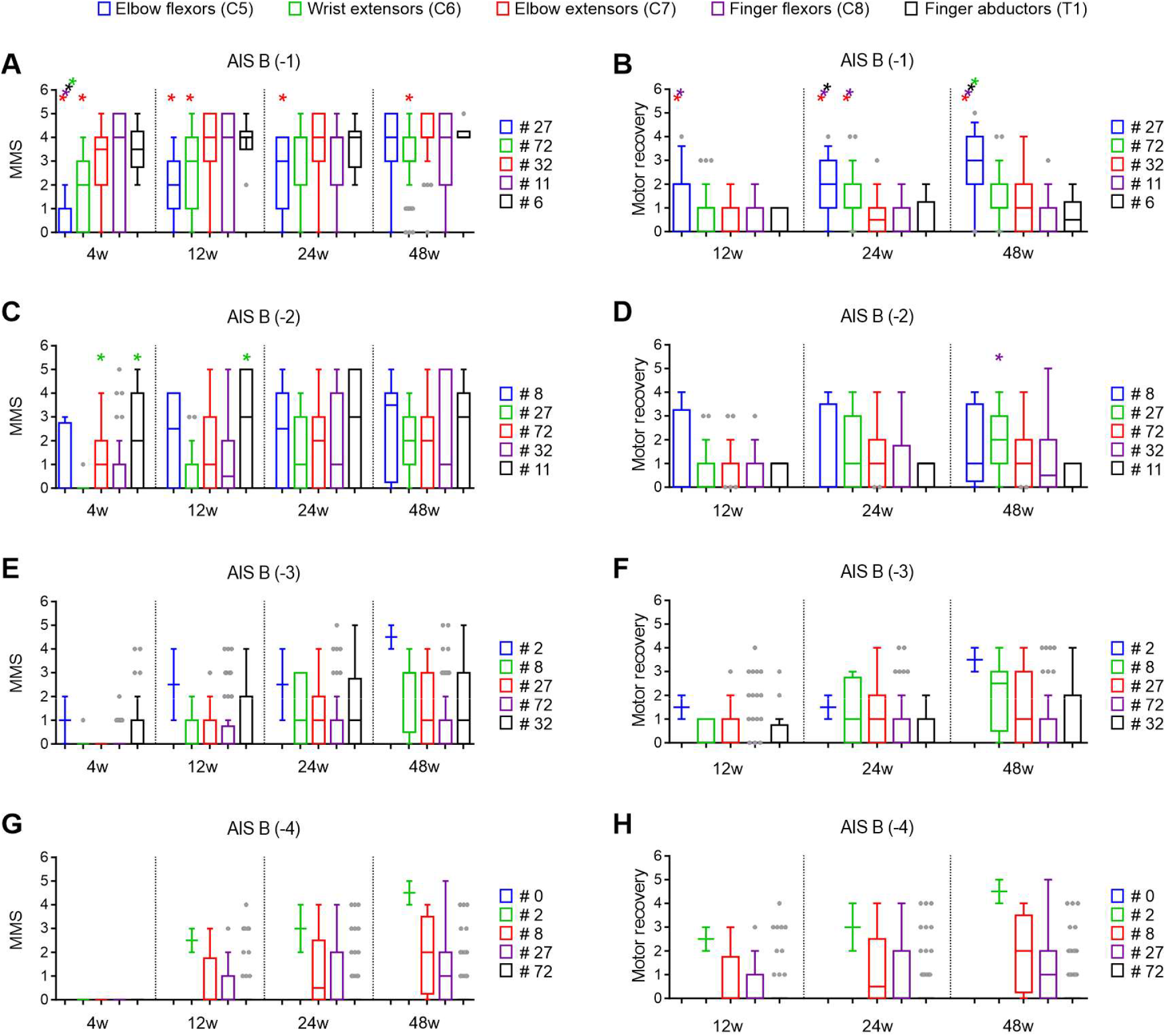
Motor recovery of muscles below the motor level of injury in AIS B individuals. (**A**) Muscles that are one level below the injury (−1) show distinct responses after the injury, the elbow flexors are more impaired. (**B**) In AIS B individuals the proximal muscles that are one level below the injury recover the most, especially the elbow flexors. (**C**) When muscles are two levels below the injury (−2) a similar trend emerges where the proximal elbow flexors and the distal wrist extensors are more affected, but also (**D**) recover to some extent. (**E-H**) No statistical analysis was conducted for muscles 3 or 4 levels (−3 or −4) below the injury because of the reduced sample size for some muscles. Nonetheless, the trends indicate a similar behavior to AIS A individuals, where distal muscles such as the finger flexors and abductors are more impaired concerning more proximal muscles. Kruskal-Wallis test with Dunn’s multiple comparisons test: *P < 0.05 relative to elbow flexors (*blue), wrist extensors (*green), elbow extensors (*red), finger flexors (*purple) or finger abductors (*black). Note: data are median (line in the middle of the box), the first quartile forms the bottom, and the third quartile forms the top of the box, whiskers represent the 5-95 percentiles; any data beyond the whiskers are shown as points (grey dots); the numbers on the right of each panel indicates the number of muscles available for each analysis. SCI = Spinal Cord Injury; AIS = American Spinal Cord Injury Association Impairment Scale; MMS = Muscle Motor Score; C = Cervical level; T = Thoracic level.

**Supplementary Figure 4.**
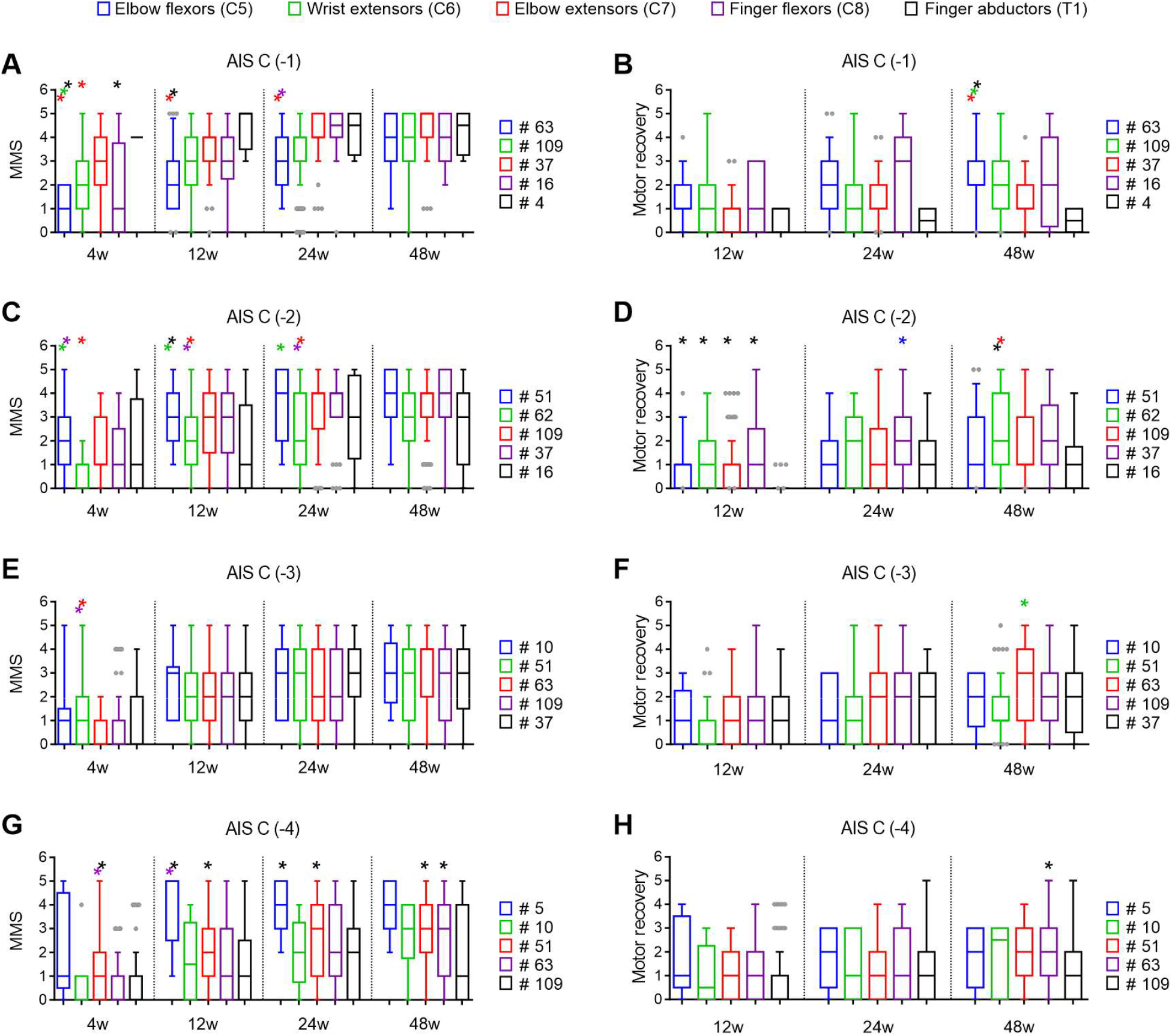
Motor recovery of muscles below the motor level of injury in AIS C individuals. (**A**) Muscles that are one level below the injury (−1) show distinct responses after the injury, the elbow flexors were more impaired and (**B**) recover to a greater extent compared to wrist extensors and finger abductors. (**C**) When muscles are 2 levels below the injury (−2) elbow flexors are less affected early after the injury, but all muscles recover to a similar grade 48 weeks post-SCI. (**D**) Early after the injury, all muscles recover more than the finger abductors. The wrist extensors show a greater recovery compared to elbow extensors and finger abductors 48 weeks post-SCI. (**E**) When muscles are 3 levels below the injury (−3) the wrist extensors are less affected early after the injury, but all muscles recover to a similar grade 48 weeks post-SCI. (**F**) Muscle strength recovery is greater for elbow extensors compared to wrist extensors. (**G**) When the lesion is distant from the muscles level (i.e., -4 levels) the proximal elbow flexors and extensors are less affected compared to finger flexors and abductors. (**H**) Muscle strength recovery is greater for the finger flexors compared to finger abductors 48 weeks post-SCI. Kruskal-Wallis test with Dunn’s multiple comparisons test: *P < 0.05 relative to elbow flexors (*blue), wrist extensors (*green), elbow extensors (*red), finger flexors (*purple) or finger abductors (*black). Note: data are median (line in the middle of the box), the first quartile forms the bottom, and the third quartile forms the top of the box, whiskers represent the 5-95 percentiles; any data beyond the whiskers are shown as points (grey dots); the numbers on the right of each panel indicates the number of muscles available for each analysis. SCI = Spinal Cord Injury; AIS = American Spinal Cord Injury Association Impairment Scale; MMS = Muscle Motor Score; C = Cervical level; T = Thoracic level.

**Supplementary Figure 5.**
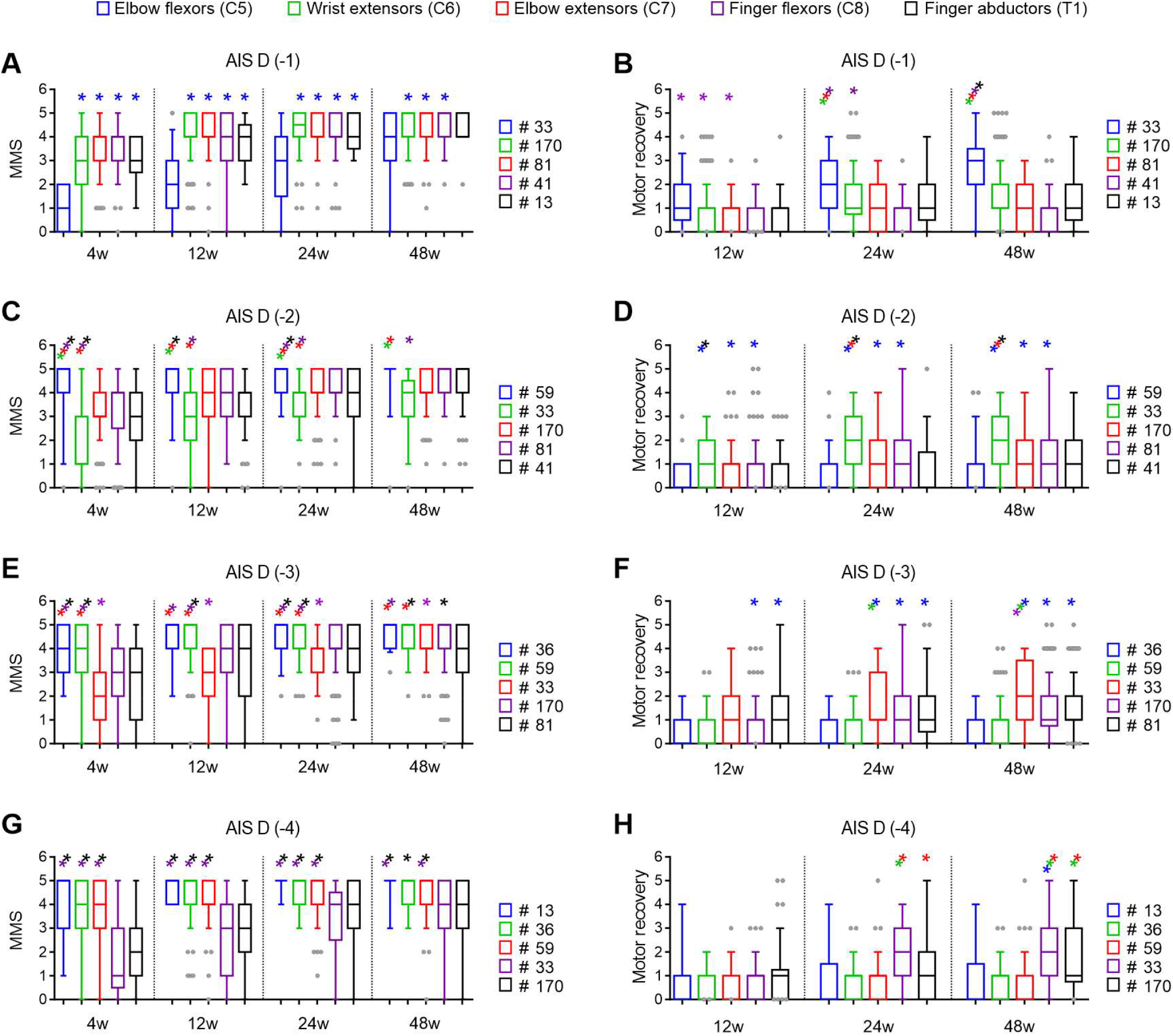
Motor recovery of muscles below the motor level of injury in AIS D individuals. (**A**) Muscles that are one level below the injury (−1) show distinct responses after the injury, the elbow flexors are more impaired compared to all other muscles. (**B**) The elbow flexors also recover the most, compared to all other muscles 48 weeks post-SCI. (**C**) If the lesion is 2 levels above the muscle level (−2), elbow flexors are less affected and wrist extensors are more affected but also (**D**) recover to a greater extent compared to elbow extensors and finger abductors (note the differences in motor score recovery concerning elbow flexors should be interpreted with caution because of ceiling effects). (**E, F**) When the SCI is 3 or 4 levels above the muscle level (−3 or -4), the distal muscles such as the finger flexors and abductors are more impaired concerning the proximal muscles. (**G, H**) These distal muscles also recover to a greater extent compared to proximal muscles (note the differences in motor score recovery concerning elbow flexors should be interpreted with caution because of ceiling effects). Kruskal-Wallis test with Dunn’s multiple comparisons test: *P < 0.05 relative to elbow flexors (*blue), wrist extensors (*green), elbow extensors (*red), finger flexors (*purple) or finger abductors (*black). Note: data are median (line in the middle of the box), the first quartile forms the bottom, and the third quartile forms the top of the box, whiskers represent the 5-95 percentiles; any data beyond the whiskers are shown as points (grey dots); the numbers on the right of each panel indicates the number of muscles available for each analysis. SCI = Spinal Cord Injury; AIS = American Spinal Cord Injury Association Impairment Scale; MMS = Muscle Motor Score; C = Cervical level; T = Thoracic level.

**Supplementary Table 1.** Demographics and baseline clinical parameters.

| Demographics |  | Full sample | Very acute | MEP sample | SSEP sample | NCS sample |
| --- | --- | --- | --- | --- | --- | --- |
| *n* |  | 748 | 440 | 203 | 313 | 280 |
| Gender |  | 599 M, 149 F | 343 M, 97 F | 171 M, 32 F | 254 M, 59 F | 228 M, 52 F |
| Age (Mean ± SD) | | 46.5 ± 18.9 | 47.1 ± 19.1 | 43.8 ± 18.8 | 44.4 ± 18.8 | 45.0 ± 19.0 |
| Baseline |  |  |  |  |  |  |
| Clinical parameters |  | Full sample | Very acute | MEP sample | SSEP sample | NCS sample |
| Neurological level<br>of injury | C1 | 22 | 24 | 4 | 7 | 7 |
|  | C2 | 45 | 23 | 4 | 8 | 9 |
|  | C3 | 95 | 62 | 28 | 40 | 38 |
|  | C4 | 279 | 168 | 81 | 120 | 109 |
|  | C5 | 189 | 88 | 46 | 83 | 70 |
|  | C6 | 72 | 33 | 25 | 38 | 31 |
|  | C7 | 33 | 7 | 14 | 15 | 14 |
|  | C8 | 13 | 4 | 1 | 2 | 2 |
|  | NT | 0 | 31 | 0 | 0 | 0 |
| AIS grade | A | 261 | 153 | 56 | 87 | 77 |
|  | B | 84 | 51 | 31 | 52 | 45 |
|  | C | 155 | 98 | 44 | 66 | 54 |
|  | D | 241 | 121 | 71 | 107 | 103 |
|  | NT | 7 | 17 | 1 | 1 | 1 |

MEP = motor evoked potential; SSEP = somatosensory evoked potential; NCS = nerve conduction studies; M = male; F = female; SD = standard deviation; C = cervical level; AIS = american spinal injury association impairment scale; NT = not tested.

**Supplementary Table 2.**
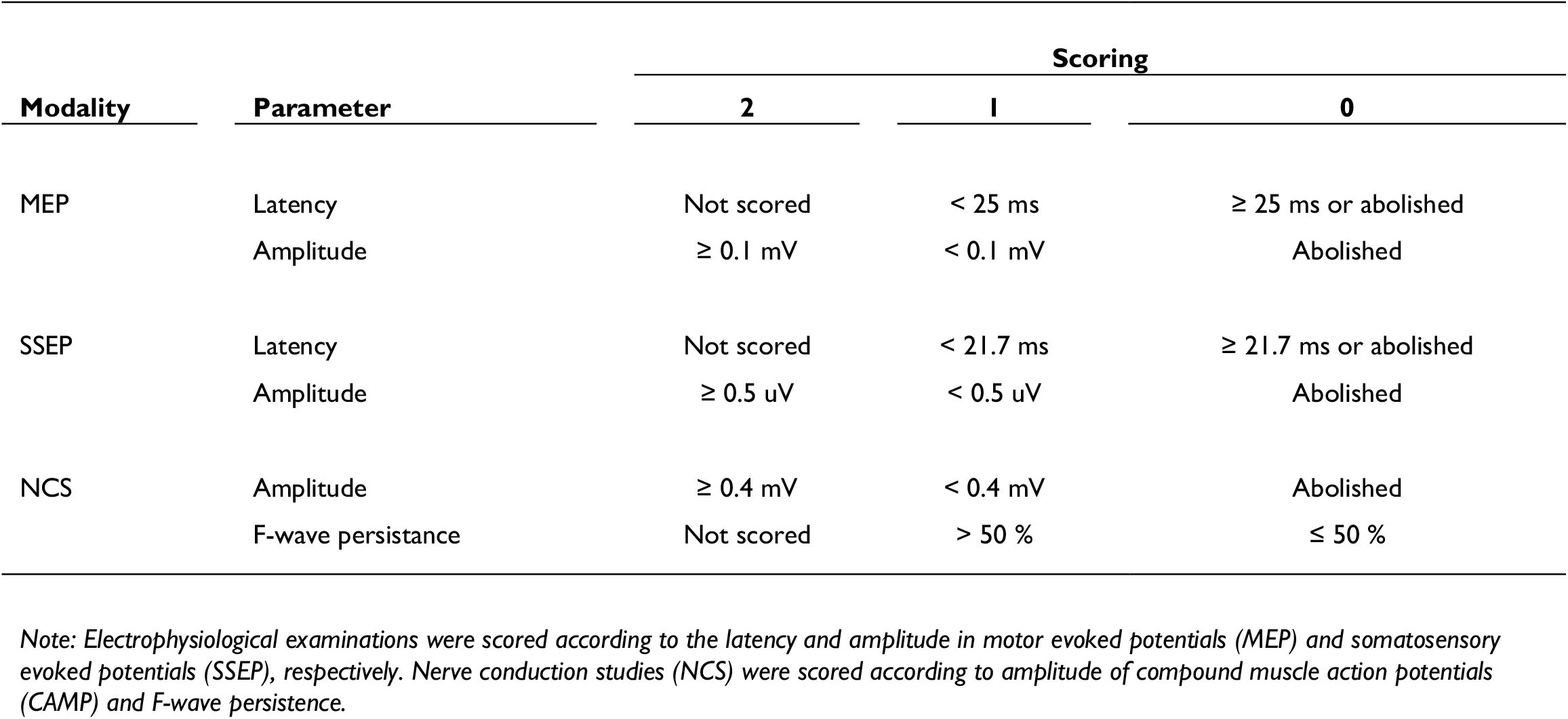
Scoring of electrophysiological examinations (Hupp, *et al.*, 2018).

**Supplementary Table 3.** Feature importance in the supervised machine learning models.

| **All AIS and muscles (Figure 4B)** | **(without muscle)** |  | **(with muscle)** |
| --- | --- | --- | --- |
| AIS | 0.24707 | AIS | 0.20526 |
| MMS | 0.25608 | Muscle | 0.1721 |
| DST | 0.19587 | MMS | 0.20074 |
| LT | 0.18199 | DST | 0.15662 |
| PP | 0.11898 | LT | 0.16093 |
|  |  | PP | 0.10435 |
| **AIS A (Figure 4C, D)** | **(without muscle)** |  | **(with muscle)** |
| MMS | 0.41186 | Muscle | 0.21887 |
| DST | 0.31671 | MMS | 0.31242 |
| LT | 0.12158 | DST | 0.23116 |
| PP | 0.14986 | LT | 0.11251 |
|  |  | PP | 0.12504 |
| **AIS B (Figure 4C, D)** | **(without muscle)** |  | **(with muscle)** |
| MMS | 0.29859 | Muscle | 0.3172 |
| DST | 0.37651 | MMS | 0.20715 |
| LT | 0.15386 | DST | 0.20443 |
| PP | 0.17104 | LT | 0.14191 |
|  |  | PP | 0.1293 |
| **AIS C (Figure 4C, D)** | **(without muscle)** |  | **(with muscle)** |
| MMS | 0.29486 | Muscle | 0.23575 |
| DST | 0.32577 | MMS | 0.24145 |
| LT | 0.18009 | DST | 0.22097 |
| PP | 0.19928 | LT | 0.14633 |
|  |  | PP | 0.1555 |
| **AIS D (Figure 4C, D)** | **(without muscle)** |  | **(with muscle)** |
| MMS | 0.35449 | Muscle | 0.25555 |
| DST | 0.35876 | MMS | 0.23421 |
| LT | 0.12353 | DST | 0.28099 |
| PP | 0.16322 | LT | 0.09585 |
|  |  | PP | 0.1334 |

*Note: Feature importance was calculated using 50% of the dataset for training and 50% for testing. AIS = American Spinal Cord Injury Association Impairment Scale; MMS = Muscle Motor Score; DST = Distance from the motor level of injury; LT = Light Touch sensation; PP = Pin Prick sensation.*

